# The Sponge Hypothesis of Sarcopenia: Functional Hyponatremia and the Liver–Muscle–Fluid Axis in Chronic Liver Disease

**DOI:** 10.64898/2025.12.06.25341772

**Authors:** Atsushi Nakamura, Takeshi Ichikawa, Keiji Okuyama

**Author notes:** Corresponding author: Atsushi Nakamura, MD, PhD, Nippon Koukan Hospital Gastroenterological Liver Disease Center, 1-2-1, Kokandori, Kawasaki-ku, Kawasaki-shi, Kanagawa 210-0852. Co-author contact details: Takeshi Ichikawa, Keiji Okuyama.

## Abstract

**Background & Aims:** Dilutional hyponatremia and sarcopenia (SP) are prognostic factors linked to portal hypertension (PHT) in chronic liver disease (CLD). Because skeletal muscle functions as the body’s largest intracellular water reservoir, early Na decline may reflect impaired muscular buffering rather than renal Na loss. This MRI-based study investigated the relationship between serum Na, SP, and PHT.

**Methods:** We retrospectively analyzed 880 CLD patients who underwent MR elastography. SP was defined as low muscle mass by MRI. Patients were categorized into five Na strata (<135, 135–136, 137–138, 139–140, ≥141 mEq/L). Individuals with Na <139 mEq/L were further stratified by ALBI score and SP to construct the liver–muscle phenotype (LMP) classification.

**Results:** SP prevalence was 28% and increased stepwise with decreasing Na levels. In non-HCC patients, Na <139 mEq/L optimally identified both SP and PHT. Liver stiffness increased progressively across SP, Na <139, and PHT (3.3→4.6 kPa, all P<0.01). Decision-tree analysis identified ALBI score (cutoff –2.16) and SP as major determinants of Na <139, producing four LMP types with graded Na decline (P<0.01). Among advanced CLD patients without ascites, worsening LMP predicted higher incidence of new-onset ascites (P<0.01). In Cox models, LMP4 (ALBI ≥–2.16 with SP) independently predicted poor prognosis (HR 3.87, P<0.01).

**Conclusions:** Functional hyponatremia (Na <139) reflects early hemodynamic stress associated with asymptomatic PHT. Given its central role in systemic water handling, distinct from the hepato–renal axis, SP may represent a modifiable target for early intervention before overt decompensation.

**Key Points:** *Significant findings of the study:*

- Functional hyponatremia (Na <139 mEq/L) and sarcopenia emerge at nearly identical liver stiffness thresholds, preceding classical portal hypertension.
- The novel Liver–Muscle Phenotype (LMP) classification identifies a high-risk group (LMP4) for fluid retention that is often underestimated by conventional MELD scores.

*What this study adds:*

- Early sodium dilution and muscle loss reflect impaired buffering by skeletal muscle—the body’s largest intracellular water reservoir—within the liver–muscle–fluid axis.
- Sarcopenia defines a reversible therapeutic window in which targeted nutrition and rehabilitation may restore fluid balance before overt retention develops.

## Introduction

Advanced chronic liver disease (ACLD) frequently presents with hyponatremia due to disturbances in electrolyte and fluid homeostasis. Sodium (Na) is essential for maintaining blood volume, blood pressure, osmotic balance, and pH, and hyponatremia has traditionally been defined as serum Na <135 mEq/L [1,2]. However, the updated Model for End-Stage Liver Disease (MELD) incorporates a broader Na range (125–137 mEq/L), highlighting that even mild sodium depletion carries prognostic significance [3]. Emerging evidence suggests that clinically relevant risk may begin even within the high-normal Na range, challenging the traditional boundary of hyponatremia.

The understanding of Na and water retention in cirrhosis has evolved over the past three decades. The hepatorenal axis model of the 1990s positioned renal vasoconstriction as the key feature of circulatory dysfunction [4], whereby portal hypertension (PHT)–induced arterial underfilling activates neurohormonal pathways, leading to renal water and Na retention and dilutional hyponatremia [2,4]. More recent refinements incorporate systemic inflammation and circulatory failure in advanced disease [5]. However, these models remain largely confined to the liver–kidney axis.

Crucially, a pathophysiological window prior to overt hyponatremia remains underexplored. In 1998, Kumar and Berl defined the normal serum sodium range as 138–142 mEq/L, emphasizing that hyponatremia develops gradually rather than abruptly and therefore requires an earlier therapeutic window [6]. In cirrhosis, Na <138–139 mEq/L has been shown to identify high-risk patients more accurately than traditional thresholds [7]. Similar associations between mildly low Na level (≤138–139 mEq/L) and adverse outcomes have been reported in heart failure [8], chronic kidney disease [9], and general oncology and geriatric populations [10]. This suggests that early Na decline represents a cross-organ phenomenon reflecting systemic fragility rather than a liver-specific event.

Sarcopenia (SP), a phenotype of malnutrition, is a well-established prognostic factor independent of hepatic function in cirrhosis [11]. However, the mechanisms through which skeletal muscle loss worsens survival, and conversely, how improving SP enhances outcomes, remain unclear. Skeletal muscle, the largest organ system in the human body by mass, consists of approximately 75% water and stores 30–40% of total body water, functioning as the principal “water-tank” in systemic fluid regulation [12,13]. In cirrhosis, muscle mass declines steadily by 2–3% per year [14], and intracellular water content is closely linked to muscle volume, strength, and frailty risk.

Based on these insights, we hypothesize that skeletal muscle functions as a systemic fluid buffer, and that loss of muscle mass represents failure of this buffering capacity—an upstream phase that may precede the decline in serum Na into the low-normal range.

## Materials and Methods

### 2.1 Study Design and Population

This retrospective cohort included 880 consecutive patients with chronic liver disease (CLD) who underwent magnetic resonance elastography (MRE) at Nippon Koukan Hospital between 2018 and 2024. Demographic data (age, sex, body mass index [BMI]), laboratory parameters, and magnetic resonance imaging (MRI) findings were collected. Blood sampling and MRI were performed within a median interval of 7 days (mean 9 ± 10 days).

Use of conventional diuretics was recorded. Patients with end-stage renal disease on dialysis, New York Heart Association class III–IV heart failure, advanced extrahepatic malignancies, or incomplete imaging data were excluded (**Supplementary Figure 1**). For patients treated with tolvaptan, laboratory and imaging data obtained before its initiation were analyzed. Hepatocellular carcinoma (HCC), a major cause of SP, was excluded from pathophysiological analyses but included in prognostic analyses. The study complied with the Declaration of Helsinki and was approved by the Institutional Review Board of Nippon Koukan Hospital (approval No. 202014). The requirement for written informed consent was waived owing to the retrospective design.

### 2.2 Disease classification, staging, and imaging measurements

All patients underwent 1.5-T MRI (GE Healthcare) after an overnight fast. MRE was used to assess liver stiffness (LS), and IDEAL-IQ sequences were used to quantify hepatic proton density fat fraction (PDFF). Patients were categorized into five Na groups: ≤134, 135–136, 137–138, 139–140, and ≥141 mEq/L. Associations between Na levels, PHT, and SP were examined. Liver fibrosis was graded using LS. Advanced chronic liver disease (ACLD) was defined as LS ≥3.0 kPa, a threshold associated with increased risk of PHT [15]. ACLD was further staged as compensated or decompensated according to the 2018 European Association for the Study of the Liver guidelines [16]. PHT was defined by the presence of at least two of the following: thrombocytopenia (<15 × 10⁴/µL), gastroesophageal varices, or ascites grade ≥1 on MRI [17,18]. Liver function and disease severity were assessed using the albumin–bilirubin (ALBI) score, Model for End-Stage Liver Disease (MELD), and MELD–Na scores [19,20].

The methodological strength of this MRI-based cohort lies in the rigorous exclusion of baseline ascites using high-resolution MRI, enabling precise identification of the clinical transition from compensated to decompensated PHT, defined as the first MRI-detectable ascites. This approach provides a clean framework to capture the moment when clinically overt ascites first appears.

### 2.3 Definition of sarcopenia

MRI was used to simultaneously quantify skeletal muscle mass, hepatic fat, and LS. SP was diagnosed using the MRI-derived paraspinal muscle index (PSMI) at the level of the superior mesenteric artery, with cut-off values of 12.62 cm²/m² in men and 9.77 cm²/m² in women [21,22]. Recent malnutrition guidelines from the American College of Gastroenterology and the American Association for the Study of Liver Diseases define disease-related sarcopenia as a phenotypic manifestation of malnutrition, primarily characterized by reduced skeletal muscle mass; muscle strength tests are considered functional and prognostic rather than diagnostic [23,24]. In line with this concept, we defined SP solely on the basis of reduced skeletal muscle mass and conceptualized muscle as a major water reservoir.

### 2.4 Liver–muscle phenotype classification

Functional hyponatremia was defined as a clinically relevant decrease in serum sodium (Na <139 mEq/L), distinct from conventional hyponatremia (Na <135 mEq/L). To explore the mechanisms and prognostic implications of this mild sodium decline, we constructed a liver–muscle phenotype (LMP) classification based on the combination of ALBI score and SP, as detailed in the Results.

### 2.5 Outcome and follow-up

Patients with ACLD were followed until liver-related death, liver transplantation, or last confirmed contact. Liver-related mortality included deaths from hepatic decompensation, variceal bleeding, spontaneous bacterial peritonitis, or HCC. Followup information was obtained from medical records, inter-institutional correspondence, and direct contact. The median follow-up period was 36 months (interquartile range [IQR] 3–87 months).

### 2.6 Statistical analysis

Statistical analyses were performed using JMP version 19.0 (SAS Institute Japan). Between-group differences were assessed using parametric or non-parametric tests, as appropriate. Variables with p<0.05 in univariable analyses and clinically relevant covariates were entered into multivariable models.

For predicting serum Na <139 mEq/L, variables identified as significant in multivariable logistic regression were subsequently entered into a classification and regression tree (CART) model, from which the LMP classification was derived. Multiple testing was controlled with the Benjamini–Hochberg false discovery rate (FDR); FDR-adjusted p<0.05 was considered significant. Final logistic regression and Cox proportional hazards models were obtained using stepwise selection. Survival was analyzed using Kaplan–Meier curves and log-rank tests, with p<0.05 regarded as statistically significant.

### 2.7 Use of AI-based tools

AI-based language tools (ChatGPT, OpenAI) were used to assist with drafting and editing; all content was reviewed and approved by the authors.

## Results

### 1. Clinical characteristics (Table 1)

Among 880 patients with CLD, 369 (42%) had ACLD. Metabolic and viral etiologies predominated, with MASLD, ArLD, and viral hepatitis together accounting for over 80% of cases (**Table 1**). ACLD patients were older, more often male, and had more advanced liver dysfunction and hemodynamic derangement, including lower serum sodium. PHT was present in 20% of the cohort, almost entirely in ACLD. Nutritional indices indicated pronounced muscle loss, with sarcopenia in 28% overall and 40% of ACLD patients, and significantly reduced PSMI in both sexes.

**Table 1.**
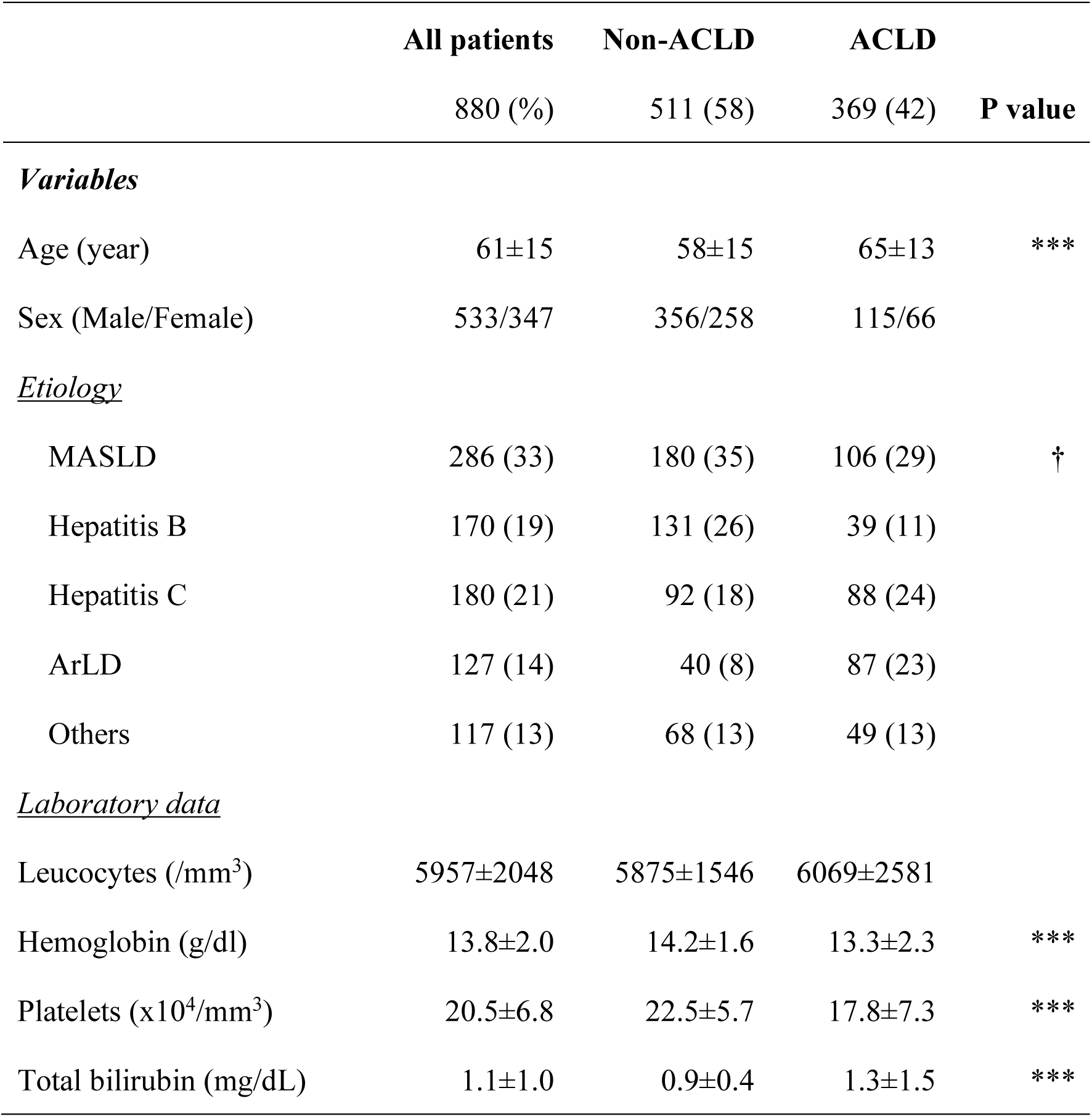
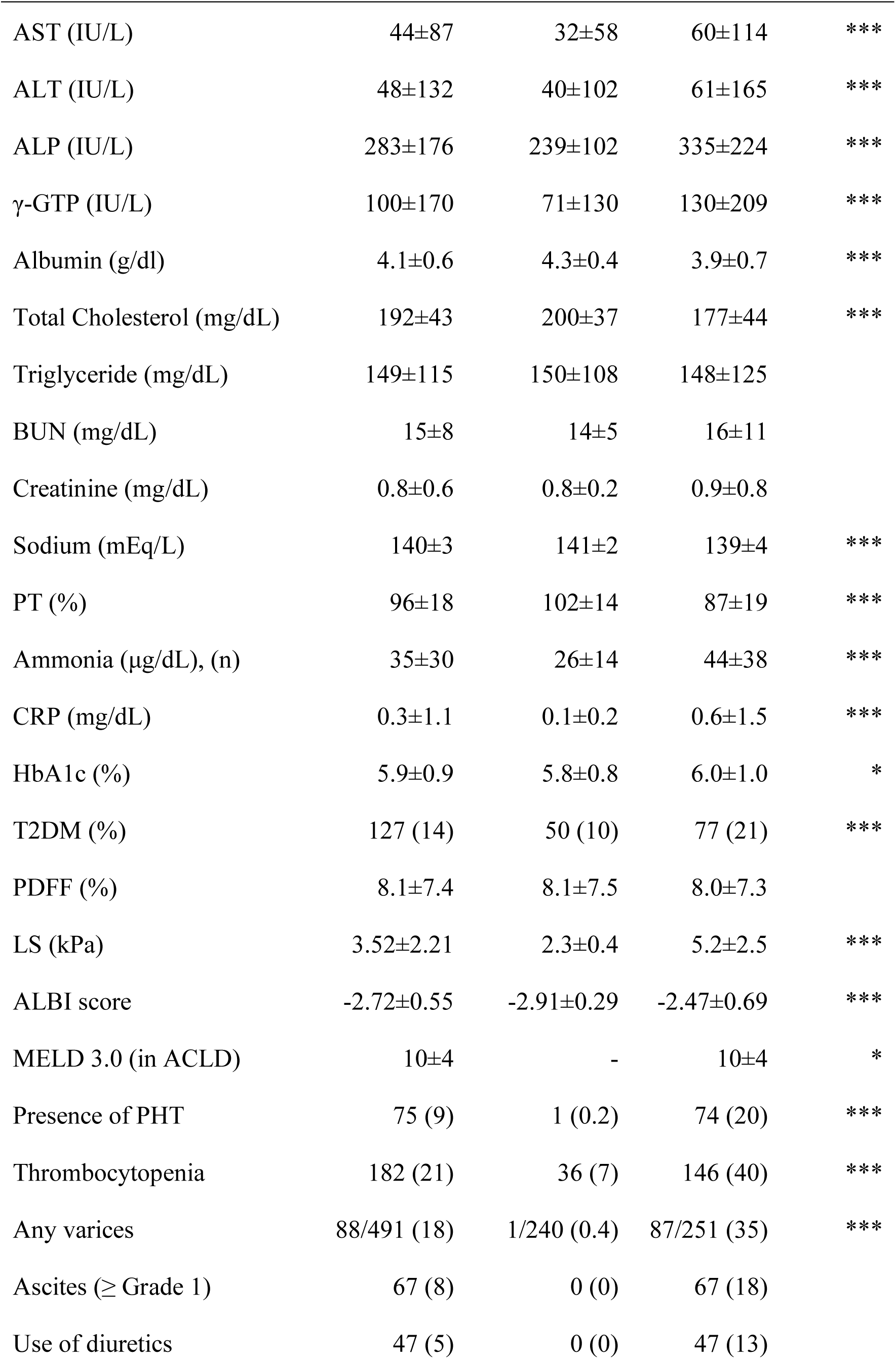
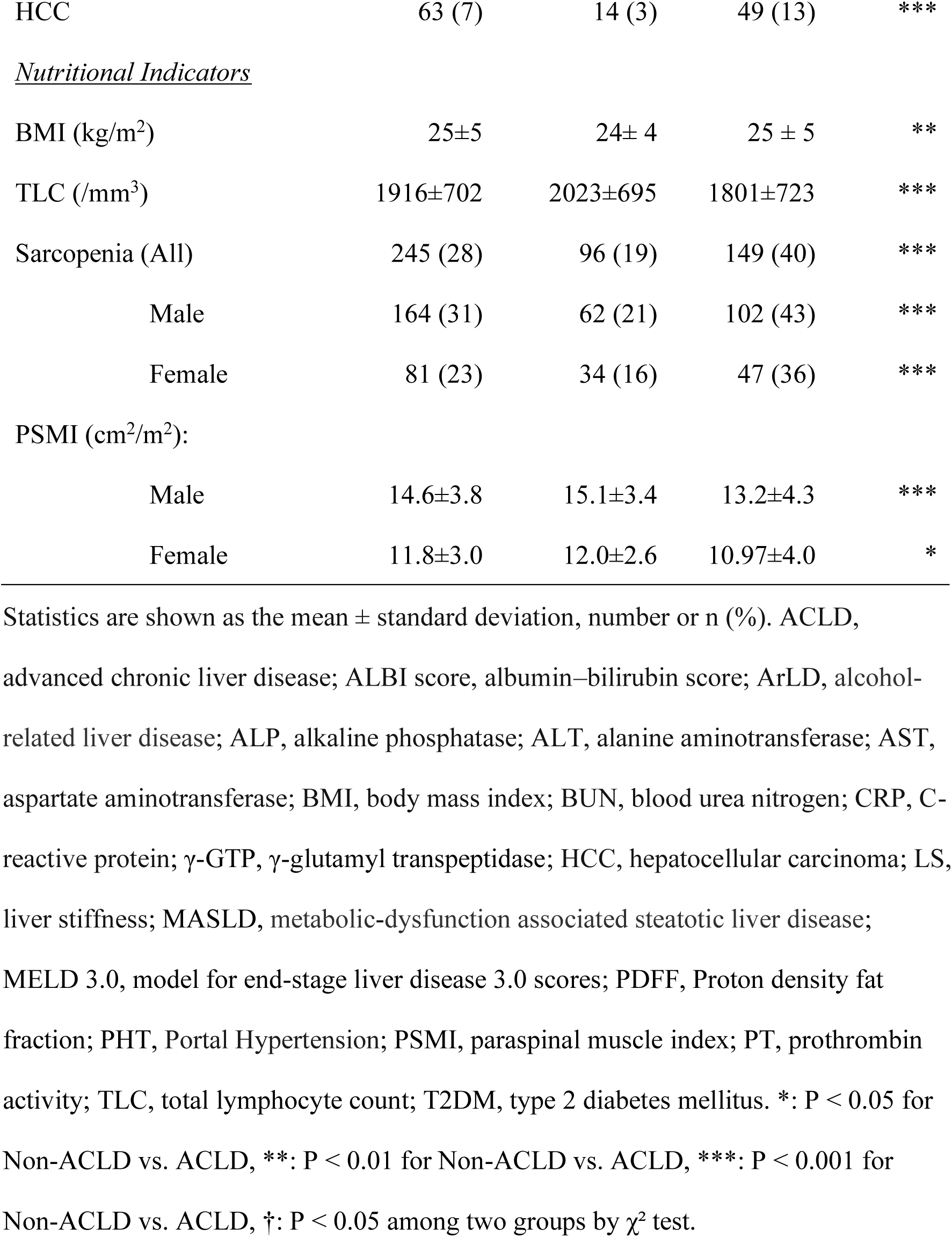
Clinical characteristics of the study population stratified by clinical stages.

### 2. Association between serum Na, LS, and SP (non-HCC cohort)

Among 822 CLD patients without HCC, lower serum Na (mEq/L) was associated with higher LS (kPa), more frequent PHT and sarcopenia, and lower muscle mass in both sexes (**Figure 1**), independent of age (**Supplementary Figure 2A**). ROC analyses identified Na <139 mEq/L as the optimal cutoff for predicting both SP and PHT (**Figure 2**), with similar performance in males and females (**Supplementary Figure 2B**). The ROC analysis identified 138 mEq/L as the optimal cutoff. Because serum Na is measured in 1-mEq/L steps, we defined functional hyponatremia as Na <139 mEq/L (i.e., ≤138 mEq/L).

**Figure 1.**
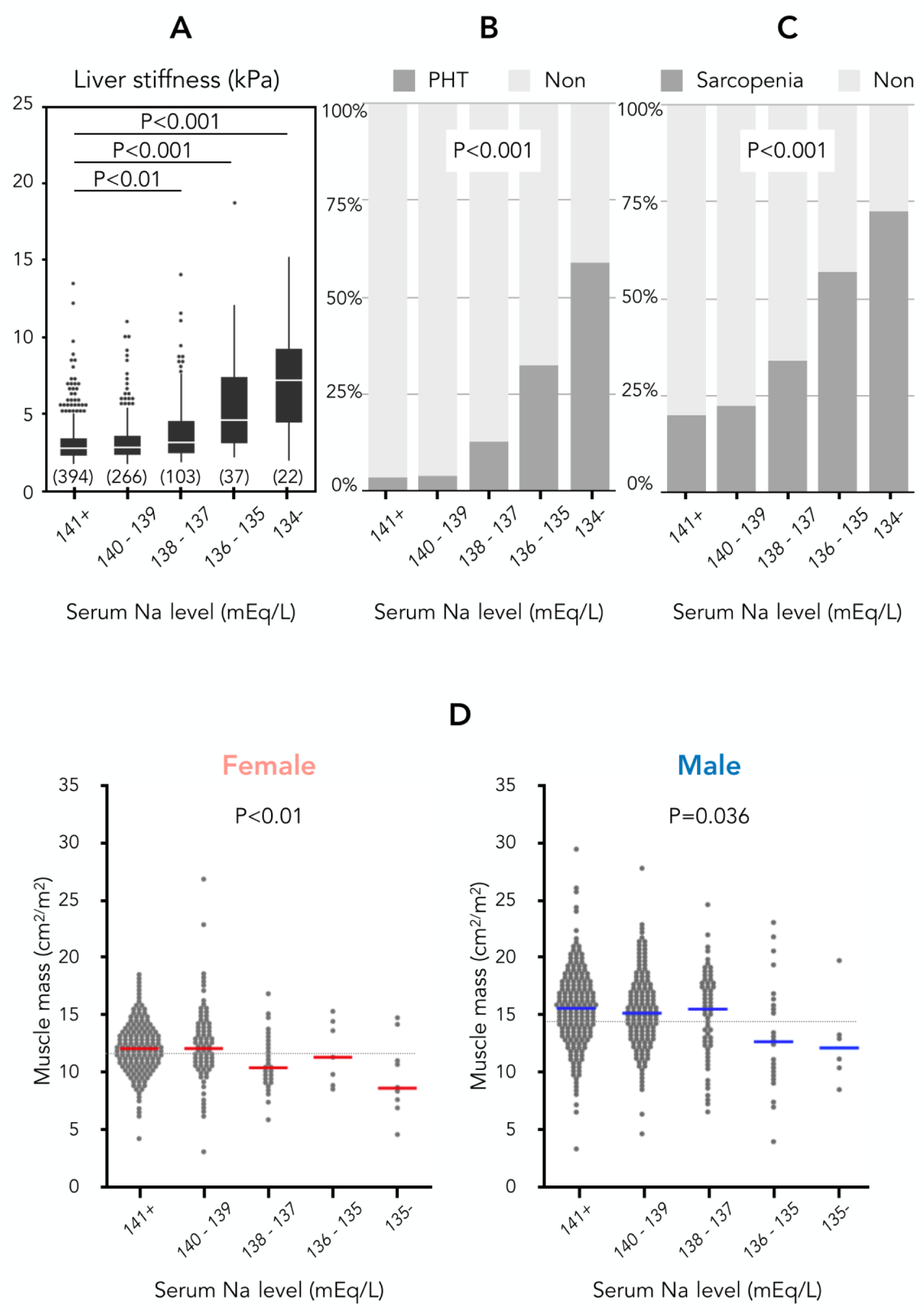
Association between Na levels and liver stiffness (LS), portal hypertension (PHT), sarcopenia (SP), and muscle mass. (A) LS across Na categories. (B) Proportion of patients with PHT. (C) Proportion of patients with SP. (D) Paraspinal muscle index (PSMI) by Na category in female (left) and male (right); bars indicate median values.

**Figure 2.**
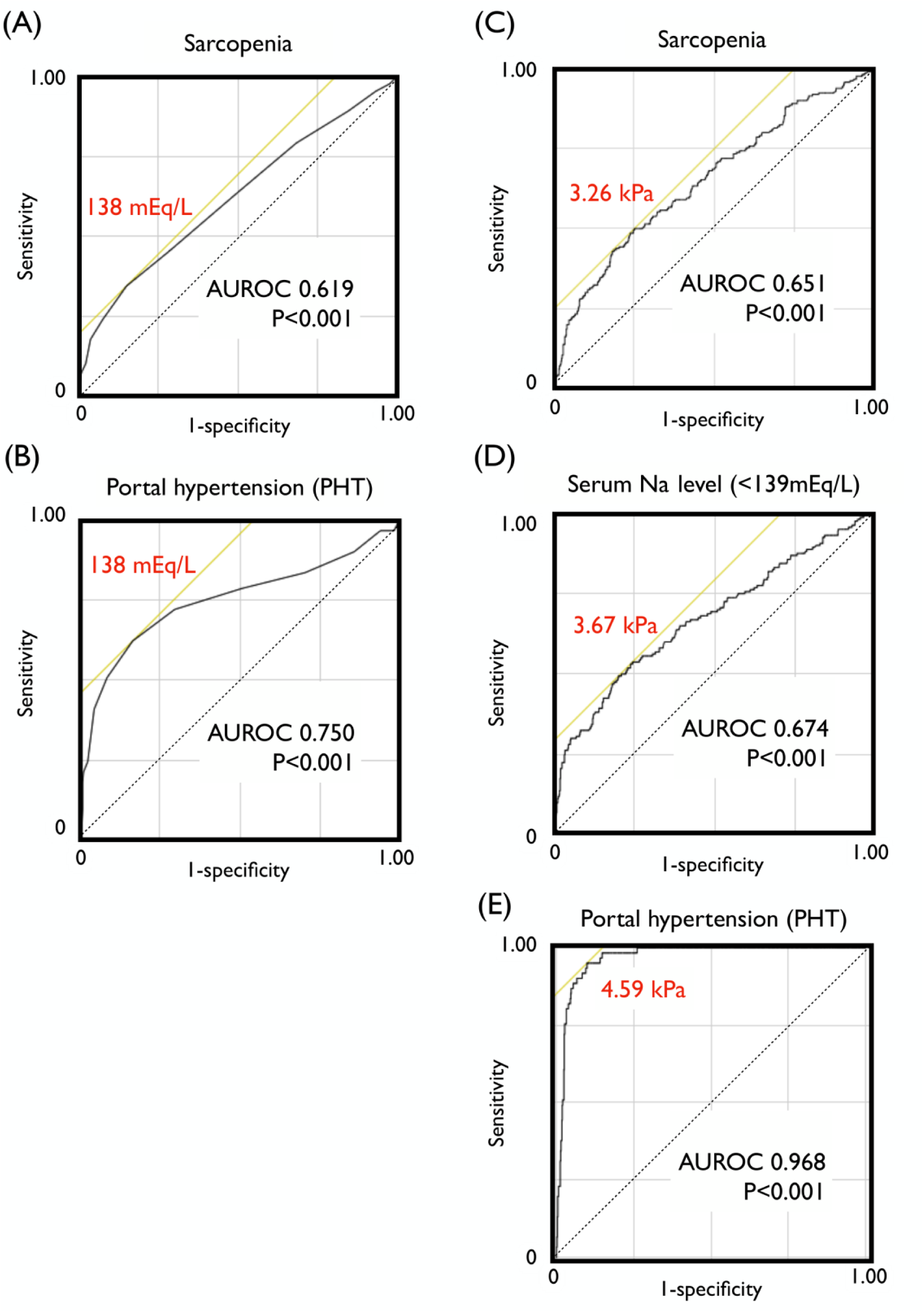
Receiver operating characteristic (ROC) analyses identifying optimal serum Na and liver stiffness (LS) thresholds associated with sarcopenia (SP) and portal hypertension (PHT). (A–B) Na (mEq/L) predicting SP and PHT. (C–E) LS (kPa) predicting SP, Na <139 mEq/L, and PHT, respectively. Optimal thresholds (Na 138 mEq/L; LS 3.26, 3.67, and 4.59 kPa) and area under ROC values are shown in each panel.

LS thresholds increased stepwise—3.26 kPa for SP, 3.67 kPa for Na <139 mEq/L, and 4.59 kPa for overt PHT—indicating that functional hyponatremia (Na <139 mEq/L) is an early marker linking fibrosis progression and SP before clinically evident PHT.

### 3. Risk Factors for Functional Hyponatremia (Na <139 mEq/L)

In the non-HCC cohort (n=822), several markers of hepatic dysfunction, PHT, inflammation, and malnutrition were associated with Na <139 mEq/L in univariable analyses (**Supplementary Table 1**). In multivariable analysis, higher ALBI score, presence of SP, male sex, higher LS, and elevated leukocyte count remained independently associated with functional hyponatremia (**Figure 3A**).

**Figure 3.**
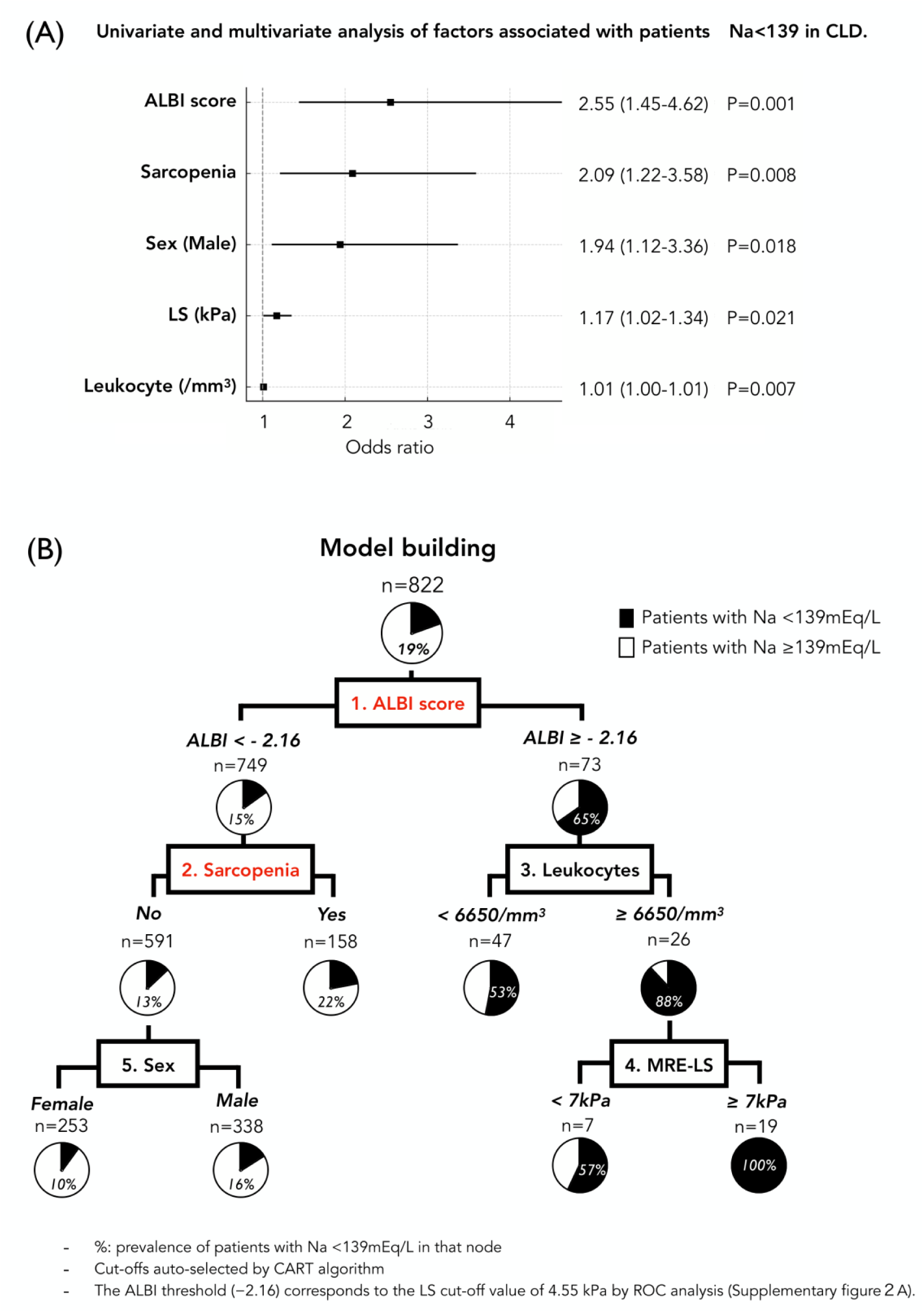
Factors associated with functional hyponatremia (Na <139 mEq/L) and decision tree analysis. **(A)** Multivariate logistic regression identifying variables independently associated with functional hyponatremia (Na <139 mEq/L) in 822 CLD patients: ALBI score, sarcopenia (SP), male sex, liver stiffness (LS), and leukocyte count. **(B)** Classification and regression tree (CART) model showing hierarchical determinants of Na <139 mEq/L. ALBI score was the primary splitter (cutoff –2.16), followed by SP, leukocyte count, sex, and LS. Percentages indicate the prevalence of Na <139 mEq/L in each terminal node.

### 4. Decision Tree Analysis (Figure 3B)

In the non-HCC cohort, CART analysis selected ALBI score as the primary splitter, with –2.16 as the optimal threshold (corresponding to 4.55 kPa on LS; **Supplementary Figure 3A**). Among patients with preserved hepatic reserve (ALBI < –2.16), SP was the next discriminator, whereas in those with ALBI ≥ –2.16, leukocyte count and then LS defined higher-risk nodes. A diuretic-adjusted model yielded a similar hierarchy (**Supplementary Figure 3B**). The final tree stratified the prevalence of functional hyponatremia from 10% in the lowest-risk leaf to 100% in the highest-risk leaf.

### 5. Liver–Muscle Phenotype (LMP) Classification (Figure 4)

Based on the CART hierarchy, we defined four liver–muscle phenotypes (LMP1–LMP4) combining ALBI score and SP status. Serum Na declined stepwise from 141 mEq/L in LMP1 to 136 mEq/L in LMP4 (overall p<0.001). CRP levels were significantly higher in LMP3 and LMP4, whereas renal function was similar across phenotypes; LMP2 patients were the oldest. Thus, the LMP framework integrates hepatic reserve and sarcopenia to capture their joint impact on Na homeostasis and systemic inflammation.

**Figure 4.**
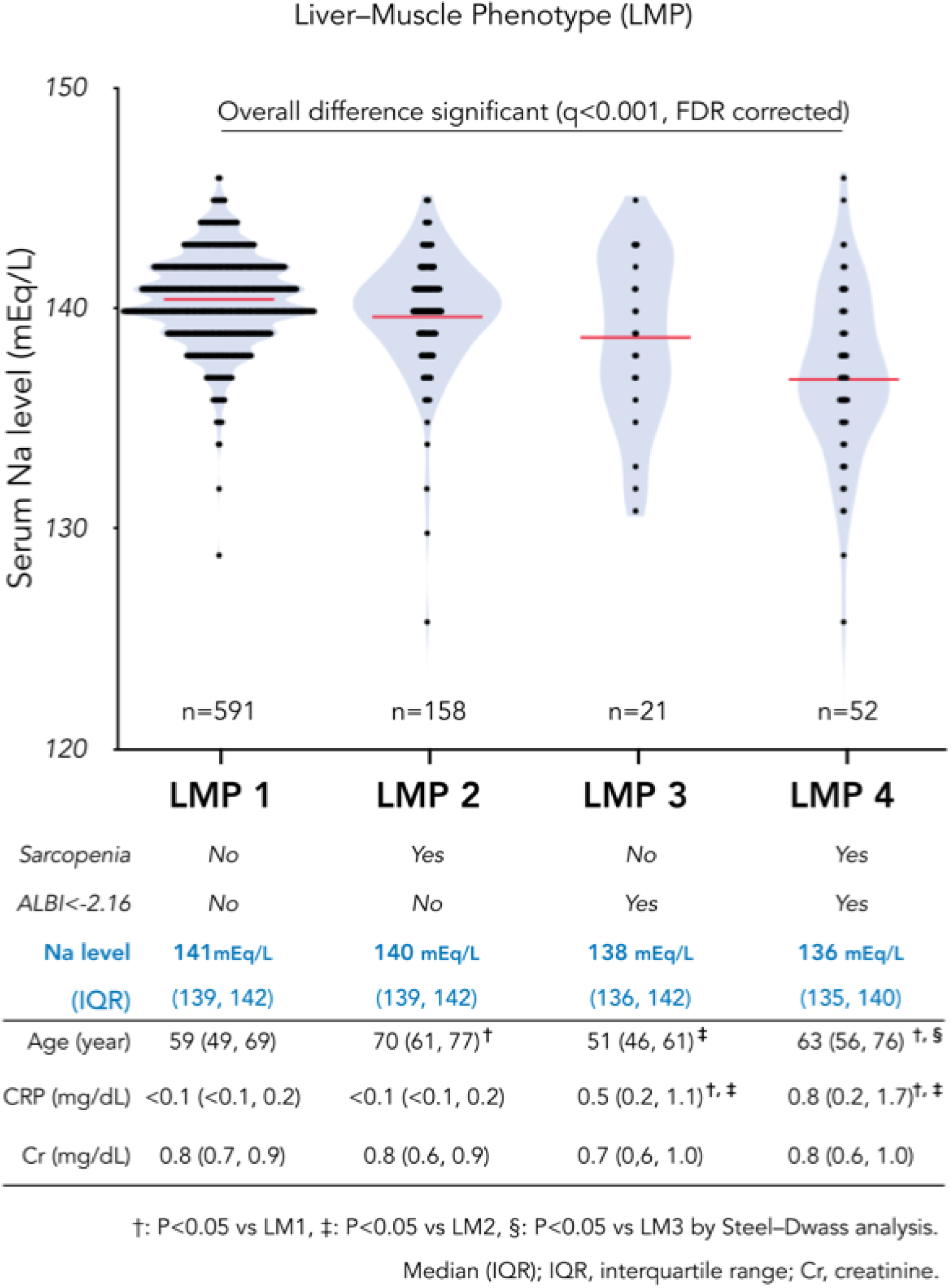
Serum Na distribution across the Liver–Muscle phenotype (LMP). Serum Na levels in four LMP phenotypes defined by hepatic reserve (ALBI < or ≥ – 2.16) and SP status; median Na declines stepwise from 141 (LMP1) to 136 mEq/L (LMP4; q<0.001). The accompanying table summarizes Na (median, IQR) and key clinical variables, with post-hoc Steel–Dwass tests for pairwise comparisons.

### 6. Prognostic impact of LMP and serum Na on new-onset ascites

Among 237 ACLD patients without baseline ascites who underwent MRI/MRE, 27 (11%) developed new-onset grade ≥1 ascites during a median 37-month follow-up. The cumulative incidence was lowest in LMP1, intermediate in LMP2/3, and highest in LMP4 (**Figure 5A**). In multivariable Cox models, LMP4 was independently associated with incident ascites compared with LMP1 and LMP2/3, whereas LMP2/3 did not differ from LMP1. Consistent with this pattern, patients with functional hyponatremia (Na <139 mEq/L) had a higher risk of ascites than those with Na ≥139 mEq/L (aHR 3.07, 95% CI 1.21–7.82; p=0.019; **Supplementary Figure 4**).

**Figure 5.**
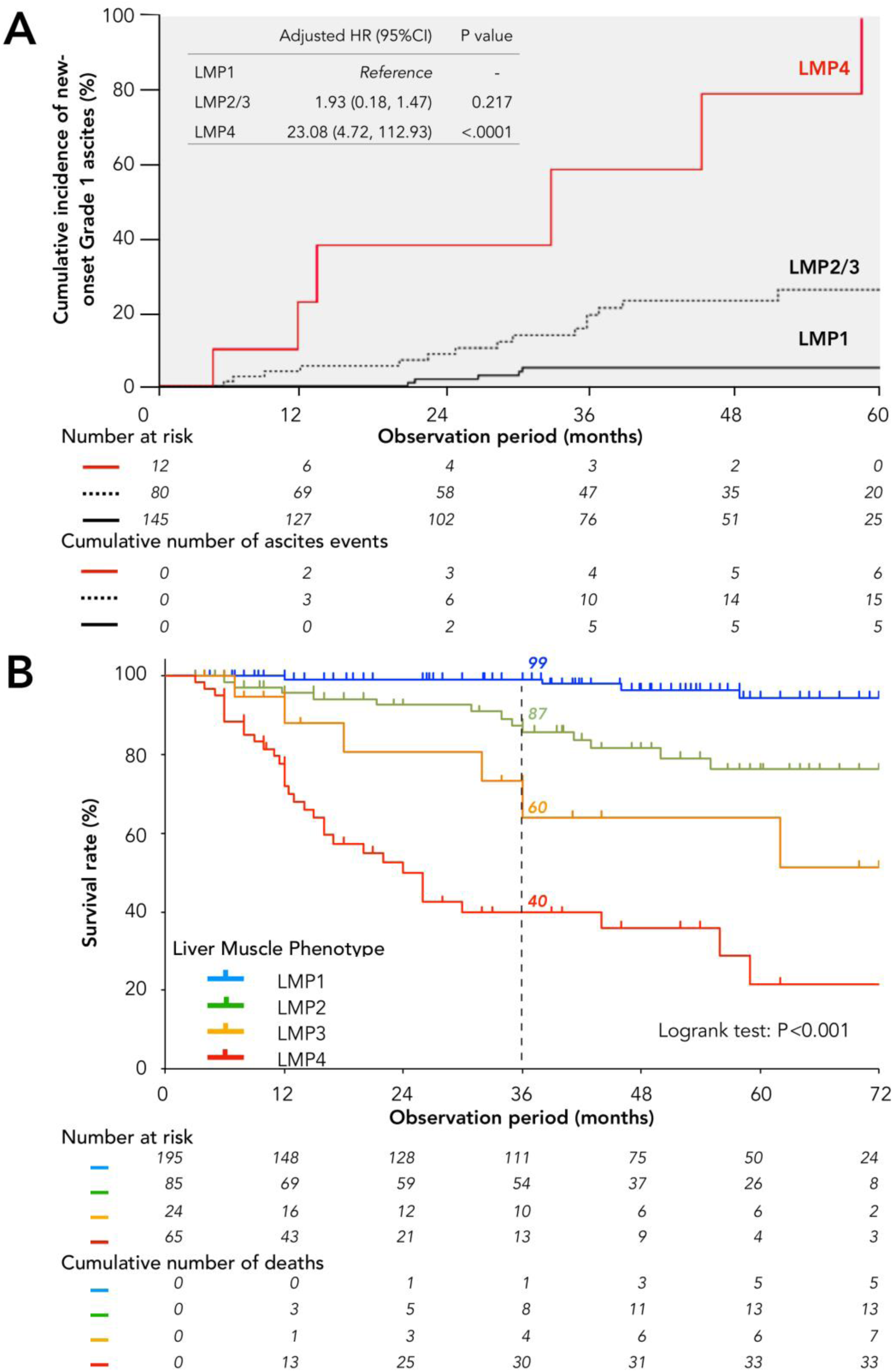
Prognostic impact of the Liver–Muscle Phenotype (LMP). (A) Cumulative incidence of new-onset grade ≥1 ascites by LMP phenotype; LMP4 shows the highest risk, whereas LMP2/3 are similar to LMP1 after adjustment for clinical covariates. Numbers at risk and cumulative events are shown below the curves. (B) Overall survival by LMP phenotype; survival declines stepwise from LMP1 to LMP4 (log-rank p<0.001). Numbers at risk and deaths are shown beneath the curves.

### 7. Prognostic impact of functional hyponatremia and LMP in ACLD

Among 369 ACLD patients (mean follow-up 35 ± 25 months; median 36 months), 58 liver-related events occurred (38 liver failures, 18 HCCs, and 2 liver transplants). Survival differed markedly across LMP stages, with the worst outcomes in LMP4, followed by LMP3 and LMP2, whereas LMP1 showed the best prognosis (**Figure 5B**). This gradient remained significant after adjustment for age, sex, LS, HCC, etiology, and diuretic use (**Table 2**). Survival also declined stepwise across sodium categories, supporting the prognostic relevance of functional hyponatremia (Na <139 mEq/L), independent of HCC status (**Supplementary Figures 6–7**). In multivariable analyses, LMP4 was a strong independent predictor of liver-related mortality alongside HCC, MELD score, and lower sodium, and remained significant even in models including MELD-Na or MELD 3.0 (HR 4.97 and 3.93, both p<0.001; **Supplementary Table 3**).

**Table 2.** Univariate and multivariate Cox proportional hazards analysis of factors associated with liver-related mortality in patients with advanced chronic liver disease (ACLD).

| **Variables** | **Univariable analysis** |  | **Multivariable analysis** |  |
| --- | --- | --- | --- | --- |
|  | HR (95% CI) | P value | HR (95% CI) | P value |
| Age (y) | 1.03 (1.00–1.05) | 0.022 |  |  |
| Sex (Male) | 2.09 (1.12–3.88) | 0.014 |  |  |
| Etiology (ArLD) | 2.01 (1.16–3.48) | 0.017 |  |  |
| ALT (IU/L) | 0.98 (0.97–0.98) | <0.001 | 0.99 (0.98–1.00) | 0.087 |
| ALP (IU/L) | 1.01 (1.00–1.01) | <0.001 |  |  |
| Total Cholesterol (mg/dL) | 0.98 (0.97–0.99) | <0.001 |  |  |
| Triglyceride (mg/dL) | 0.99 (0.98–0.99) | <0.001 | 0.99 (0.99–1.00) | 0.147 |
| CRP (mg/dL) | 1.31 (1.18–1.42) | <0.001 |  |  |
| Hemoglobin (g/dl) | 0.72 (0.65–0.80) | <0.001 |  |  |
| Platelets ($\times 10^4/\text{mm}^3$) | 0.87 (0.83–0.91) | <0.001 | | |
| Lymphopenia (<<br>1500/$\text{mm}^3$) | 4.42 (2.52–7.73) | <0.001 | | |
| Sodium (mEq/L) | 0.85 (0.82–0.89) | <0.001 | **0.90 (0.85–0.96)** | **0.004** |
| LS (kPa) | 1.23 (1.15–1.30) | <0.001 |  |  |
| PDFF (%) | 0.88 (0.81–0.94) | <0.001 |  |  |
| ALBI score | 4.50 (3.23–6.29) | <0.001 |  |  |
| MELD score | 1.20 (1.13–1.25) | <0.001 | **1.08 (1.01–1.14)** | **0.021** |
| Decompensated ACLD | 7.09 (4.15–12.1) | <0.001 |  |  |
| Presence of HCC | 5.13 (3.03–8.67) | <0.001 | **5.50 (2.96–10.22)** | **<.001** |
| Sarcopenia | 6.10 (3.16–11.79) | <0.001 |  |  |
| LMP 4 (vs. LMP 1-3) | 10.19 (5.93–17.49) | <0.001 | **3.87 (1.93–7.75)** | **<.001** |

HR, Hazard Ratio; CI, Confidence Interval; ArLD, Alcohol-related Liver Disease; ALT, Alanine Aminotransferase; ALP, Alkaline Phosphatase; CRP, C-reactive Protein; LS, Liver Stiffness; PDFF, Proton Density Fat Fraction; ALBI score, Albumin–Bilirubin score; MELD score, Model for End-stage Liver Disease score; ACLD, Advanced Chronic Liver Disease; HCC, Hepatocellular Carcinoma; LMP, Liver-Muscle Phenotype.

### 8. Associations between LMP and liver prognostic scores (ALBI and MELD 3.0)

In ACLD, ALBI and MELD 3.0 showed similar score distributions in LMP3 and LMP4, indicating that these conventional scores did not distinguish the highest-risk phenotype (**Figures 6A–B**). HCC was slightly more frequent in LMP4 than in LMP3. Compared with LMP3, LMP4 patients were older and displayed features of catabolic decline—lower BMI, hemoglobin, albumin, and total cholesterol and higher CRP—despite comparable liver prognostic scores (**Supplementary Table 4**). In multivariable logistic regression, higher CRP and BUN and lower BMI independently characterized LMP4 (**Figure 6C**), indicating that LMP4 reflects an inflammation-driven chronic disease state rather than simple hepatic dysfunction.

**Figure 6.**
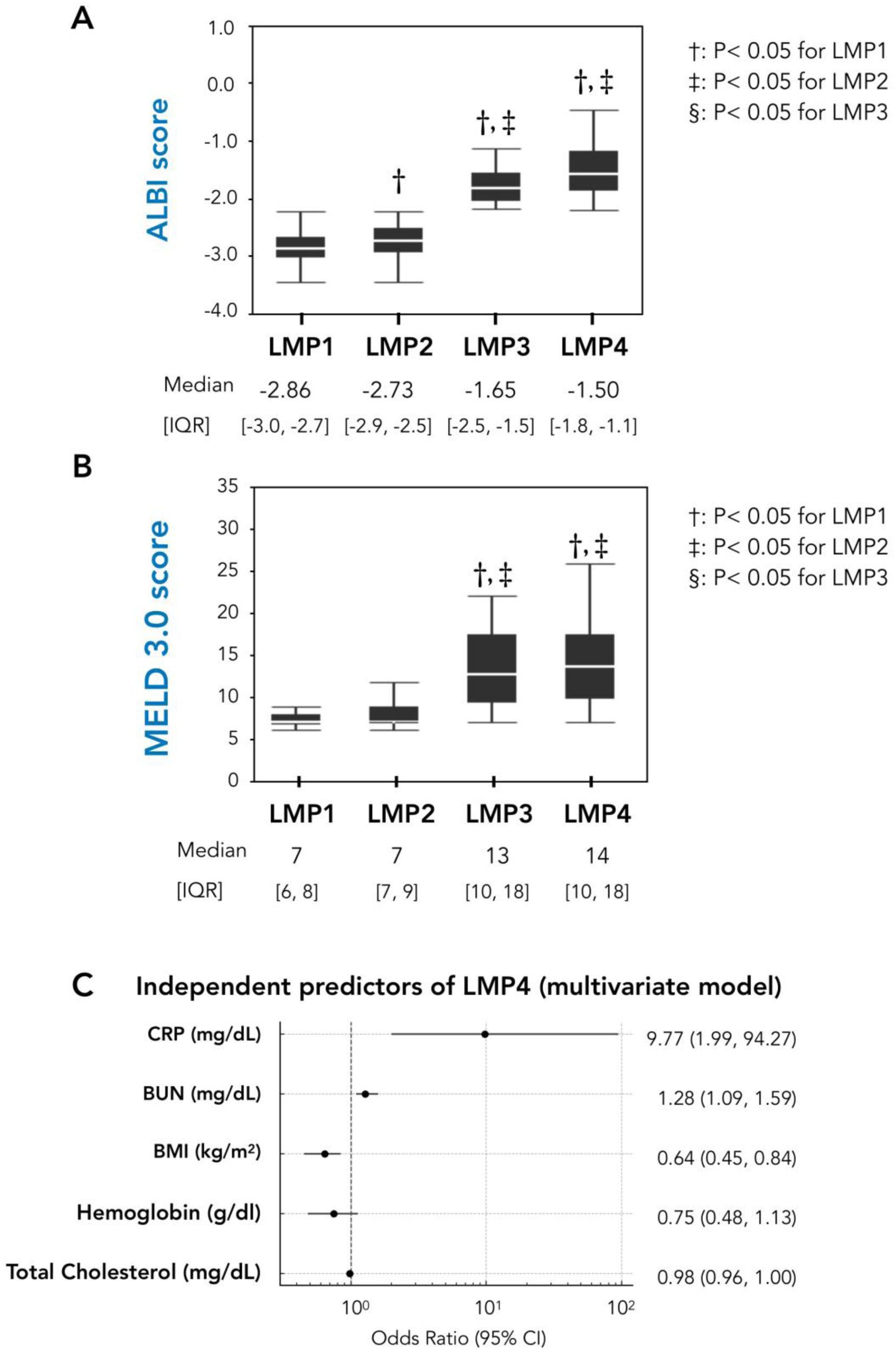
Hepatic severity scores and independent predictors of the LMP4 phenotype. (A–B) ALBI and MELD 3.0 scores across LMP1–LMP4; both worsen from LMP1 to LMP4 but do not differ between LMP3 and LMP4. Values are median (IQR); pairwise comparisons use Steel–Dwass tests († p<0.05 vs LMP1; ‡ vs LMP2; § vs LMP3). (C) Multivariable logistic regression showing higher CRP and BUN and lower BMI as independent characteristics of LMP4; odds ratios and 95% confidence intervals are shown.

## Discussion

Skeletal muscle contains approximately 70–75% intracellular water, making it the largest reservoir of intracellular fluid in the human body. MRI quantifies muscle volume rather than water content directly, but reductions in MRI-derived muscle mass inevitably represent a proportional loss of intracellular water. Bioelectrical impedance analysis (BIA), which estimates lean mass based on total body water, also underscores the fluid-rich composition of skeletal muscle. Because muscle cell volume is largely maintained by intracellular water, a reduction in intracellular fluid leads to cellular shrinkage that appears as a decrease in muscle cross-sectional area on MRI. Thus, SP can be interpreted as a reduction in the body’s intracellular water reserve.

In this context, our multimodal MRI study yielded two key findings: first, that even mild Na depletion (Na <139 mEq/L) carries clinical significance; and second, that muscle loss may contribute to abnormalities in Na and water homeostasis beyond the traditional liver–kidney axis. We initially hypothesized that SP might directly contribute to dilutional hyponatremia in ACLD. However, our findings indicated a more physiologically coherent interpretation. The relevance of SP in this setting lies not in the magnitude of its statistical association with Na dysregulation (OR 2.09), but in its early appearance alongside functional hyponatremia. Notably, both emerged at nearly identical LS thresholds and well before overt PHT, positioning them as early markers of hemodynamic stress (**Figures 1 and 2**). This co-emergence identifies a potentially reversible stage of disease progression in which targeted interventions may remain effective.

These findings suggest that SP serves not merely as a prognostic correlate but as an early pathophysiological signal marking the transition from physiological compensation to emerging circulatory stress.

### 1. Clinical Significance of Functional Hyponatremia

We defined “functional hyponatremia” as serum sodium <139 mEq/L—a value within the conventional normal range but upstream of traditional hyponatremia, potentially indicating early dysregulation of water–Na homeostasis. At this level, three pathophysiological processes begin to converge: (1) reduced intracellular water buffering capacity due to skeletal muscle loss, (2) visceral vasodilation and early effective arterial underfilling associated with portal hypertension, and (3) non-osmotic release of antidiuretic hormone preceding full activation of the renin–angiotensin–aldosterone system. Collectively, these changes characterize a subtle pre-compensatory state in which mild sodium depletion functions as an early, quantifiable signal of emerging hemodynamic stress.

This concept aligns with the physiological framework proposed by Kumar and Berl, who defined 138–142 mEq/L as the normal sodium range [6]. Importantly, the term “functional” reflects that these Na concentrations, while numerically normal, may already exert physiological effects characteristic of hyponatremia. In our cohort, functional hyponatremia was associated with a markedly elevated risk of new-onset ascites (adjusted hazard ratio 3.07, **Supplementary Figure 4**), supporting its role as an early indicator of circulatory decompensation within a potentially reversible therapeutic window.

### 2. Clinical Value of the LMP Classification System

The LMP framework—based on ALBI score and SP—revealed a clear physiological gradient that was not apparent from conventional laboratory parameters. Serum Na declined stepwise from 141 to 136 mEq/L across stages, consistent with progressive impairment of free-water handling, whereas demographic factors, BMI, and routine liver biochemistry remained heterogeneous (**Supplementary Table 2**). Hemoglobin decreased in parallel from 14.2 to 10.8 g/dL, reflecting early plasma volume expansion and mild circulatory or neurohormonal stress, further influenced by splenic sequestration as suggested by thrombocytopenia [1,2]. Thus, Na reflects free-water balance, whereas hemoglobin reflects dilutional expansion of circulating plasma volume. These changes emerge during an early hemodynamic phase preceding overt circulatory dysfunction and may therefore remain reversible. In other words, the LMP classification captures not merely Na concentration, but the accumulating diluting fluid—the body’s rising “water level.”

The prognostic strength of LMP4 derives from the convergence of two fundamental processes: reduced intracellular water–buffering capacity due to SP and impaired hepatic synthetic function (ALBI score ≥ –2.16). Notably, this ALBI threshold closely corresponded to the LS value that predicted portal hypertension on ROC analysis, supporting its physiological relevance. Together, these alterations promote redistribution of fluid from the intracellular to the extracellular compartment, manifesting as functional hyponatremia and dilutional anemia. LMP4 also identified a catabolic, inflammation-associated phenotype characterized by elevated CRP, lower BMI, hemoglobin, and cholesterol, capturing a transitional phase that precedes irreversible decompensation.

A more linear decline in both Na and hemoglobin than expected from dilution alone suggests that additional hemodynamic or metabolic factors may contribute, a question that warrants further investigation.

### 3. Clinical Implications in the Context of MELD and Prognostic Scoring

The present findings refine the interpretation of MELD-based prognostic assessment in ACLD. Although MELD-Na and MELD 3.0 incorporate key elements of circulatory dysfunction—serum creatinine, bilirubin, INR, and the presence of HCC—these scores were unable to distinguish between LMP3 and LMP4 despite their markedly different prognoses. This limitation reflects two structural vulnerabilities in MELD-based systems. First, creatinine-based eGFR overestimates true GFR in nearly half of cirrhotic patients [25], and low muscle mass is a major determinant of this overestimation. The median creatinine level in LMP4 (0.77 mg/dL) appeared “normal,” but reduced fat-free mass likely masked early declines in renal filtration. Second, bilirubin production depends largely on hemoglobin-derived heme catabolism [26–28], a relationship reproduced in our cohort (**Supplementary Figure 8**). LMP4 showed significantly lower hemoglobin than LMP3 (**Supplementary Table 4**), and bilirubin levels were paradoxically lower despite more advanced circulatory dysfunction—consistent with evidence that reductions in red cell mass directly decrease bilirubin generation independent of hepatic clearance.

Importantly, SP is strongly associated with anemia in cirrhosis [29], implying that SP may concurrently lower creatinine and modestly reduce bilirubin. These effects are particularly relevant in patients with *intermediate MELD ranges*, where small shifts in creatinine or bilirubin can meaningfully alter calculated scores. Consequently, SP may lead to a mild underestimation of disease severity by MELD-Na and MELD 3.0 in this physiological window, even though patients in LMP4 demonstrate substantially worse outcomes. These findings suggest that LMP may provide complementary information to MELD-based assessments, particularly when MELD scores fall within the intermediate range.

### 4. Toward “Hepatic Rehabilitation”: A Future Direction

Cardiac rehabilitation improves outcomes in heart failure, with peripheral muscle adaptation recognized as one contributing mechanism [30]. Drawing on this precedent, our findings suggest a potentially analogous role for skeletal muscle in ACLD: as a physiological sponge that buffers fluid shifts, its loss may precipitate early circulatory dysfunction. Unlike conventional rehabilitation aimed at restoring lost function, “hepatic rehabilitation” would thus focus on preserving this buffering capacity rather than achieving gains in muscle mass or strength per se.

This concept is supported by the cell swelling theory, in which cellular hydration acts as an anabolic signal promoting protein synthesis, whereas cell shrinkage triggers catabolism [31]. Resistance training has been shown to chronically increase intracellular water content, providing a mechanistic rationale for exercise-based interventions aimed at maintaining the intracellular fluid reservoir [32].

## Limitations

This study has several limitations.

1. It was a single-center retrospective study conducted in an Asian population, and the findings require external and prospective validation in diverse cohorts.
2. Because MRI-based analyses were limited to patients who underwent MRE, individuals with decompensated ACLD or severe complications were underrepresented. This selection bias may partially explain the discrepancy between MELD and LMP distributions and the relatively small number of patients with LMP3 or true hyponatremia.
3. This study represents a cross-sectional, hypothesis-generating pathophysiological model; therefore, no causal relationship can be inferred between serum sodium concentration and SP. Indeed, we propose that these phenomena are not cause and effect, but rather parallel manifestations of the same fluid redistribution—SP reflecting intracellular contraction and hyponatremia reflecting extracellular dilution. However, our cross-sectional data cannot directly demonstrate this compartmental shift, and longitudinal validation is required.
4. This study does not establish that hyponatremia in cirrhosis is purely dilutional, and the interpretation presented here should be considered a pathophysiological hypothesis requiring further investigation.

## Conclusions

Our findings integrate and extend two seminal frameworks that have shaped hepatorenal physiology for three decades: Arroyo’s hepatorenal axis and the Kumar–Berl concept of conservative hyponatremia. This study reframes SP from a secondary prognostic indicator to an early pathophysiological component of the liver–muscle–fluid axis, identifying it as a potential disease-modifying target. Using multimodal MRI biomarkers, we demonstrated that skeletal muscle loss emerges earlier than both functional hyponatremia and overt PHT, indicating that reduced intracellular water-buffering capacity may represent the first measurable signal of emerging circulatory stress. Low sodium and SP functioned as twin indicators positioned at opposite ends of a physiological “fluid pendulum,” in which intracellular water loss within skeletal muscle corresponded to extracellular dilution within the vascular compartment.

The LMP classification delineated four physiological phases characterized by progressive sodium dilution, extracellular fluid predominance, and a metabolic–inflammatory shift. Notably, LMP4—defined by concomitant hepatic dysfunction and SP—identified a high-risk phenotype that was not adequately captured by MELD-based systems. The nonlinear declines in sodium and hemoglobin observed exclusively within this phenotype support a mechanism of functional dilution driven by increased plasma volume and diminished intracellular water storage. Recognition of this liver–muscle–fluid axis highlights a potentially reversible window before overt decompensation. Prospective and mechanistic studies are required to determine whether preserving muscle mass and maintaining hepatic reserve can modify disease trajectory and delay clinical deterioration.

Ultimately, skeletal muscle is not merely a motor organ but the body’s largest intracellular water-buffering reservoir, functioning as a physiological “sponge.” Its dysfunction may contribute directly to the deteriorating prognosis of patients with cirrhosis.

## Data Availability

The datasets generated and analyzed during the current study are not publicly
available due to institutional restrictions on patient privacy, but de-identified
data are available from the corresponding author upon reasonable request.

## Acknowledgments

None.

## Author contributions

AN,TI, KO: study concept and design; AN,TI, KO: patient recruitment and characterisation; AN,TI, KO: data acquisition; AN,TI, KO: data analysis; AN,TI, KO: article drafting. All authors provided input and critical revision and approved the final version.

