## Supplementary Material for "The Sponge Hypothesis of Sarcopenia: Functional Hyponatremia and the Liver–Muscle–Fluid Axis in Chronic Liver Disease"

1. Methods

- Methods (1)–(5)

2. Supplementary Information

- Tables 1–4
- Figures 1–8

**1.1 Disease Classification, Staging, and Imaging Measurements**

Steatotic liver disease (SLD) was diagnosed based on imaging findings (hepatic and renal contrast on abdominal ultrasonography; liver/spleen ratio < 0.9 on abdominal computed tomography (CT); proton density fat fraction measured by magnetic resonance imaging (MRI), > 5.2%[1]. Additionally, alcohol-related liver disease was diagnosed in accordance with the diagnostic criteria of the Japanese Society for Biomedical Research on Alcohol [2], and metabolic dysfunction-associated steatotic liver disease (MASLD) was diagnosed according to the new diagnostic criteria[3]. Hepatocellular carcinoma (HCC) was diagnosed according to the guidelines as tumors marked in the arterial phase on contrast-enhanced CT and showing washout in the portal or delayed phase[4,5]. In the case of gadoxetate disodium (EOB) contrast-enhanced MRI, tumors were also defined as those that stained in the early phase and showed washout in the portal venous phase.

**1.2 Diagnostic criteria for advanced CLD (ACLD), and procedures for diagnosing the severity of hepatic reserve dysfunction:** The diagnosis of ACLD in this study was based on LS ≥ 3kPa on magnetic resonance elastography (MRE) in accordance with the practice guidance by the American Association for the Study of Liver Diseases (AASLD)[6]. Liver reserve function impairment was evaluated using the albumin-bilirubin (ALBI) score, and disease severity was categorized based on ALBI grade

(grade 1 to 3) and the Model for End-Stage Liver Disease–sodium (MELD-Na) score[7,8].

All patients underwent liver MRI after fasting for more than 12 hours, using a 1.5-T whole-body scanner (SIGNA Voyager XT 1.5T; GE Healthcare, Tokyo, Japan). The imaging protocol included axial and coronal T1- and T2-weighted images with fat suppression, diffusion-weighted imaging, and magnetic resonance elastography (MRE) using 60 Hz acoustic vibration delivered via a pneumatic driver positioned over the right thorax. Liver stiffness (LS) was measured by MRE. A 19 cm passive pneumatic driver connected to an acoustic waveform generator transmitted 60 Hz vibrations to the right thorax at the xiphoid level. Wave propagation was captured using a gradient-echo sequence, and the region of interest (ROI) was carefully placed in the right hepatic lobe, avoiding large vessels, bile ducts, gallbladder, tumors, and imaging artifacts. LS values (in kPa) were assessed by experienced radiologists. Intrahepatic fat content was quantified using the IDEAL IQ sequence to calculate proton density fat fraction (PDFF), expressed as a percentage. All imaging-derived quantitative measurements were conducted by a single trained radiologist with expertise in clinical hepatology trials. Both LS and PDFF values were used in subsequent analyses as quantitative indicators of liver fibrosis and steatosis, respectively.

### 1.3 MRI-Based Quantification and Diagnostic Criteria

・**Sarcopenia (SP).** Skeletal muscle mass was quantified on 1.5-T MRI as previously described [9,10]. Paraspinal muscle area (PSMA) was traced on T2-weighted single-shot fast spin-echo images at the level of the superior mesenteric artery (SMA). Bilateral erector spinae, multifidus, quadratus lumborum, and spinalis muscles were summed and height-adjusted to yield the paraspinal muscle index (PSMI, $cm^2/m^2$). Sex-specific cut-offs (male < 12.62 $cm^2/m^2$, female < 9.77 $cm^2/m^2$) identified MP. Repeatability was not re-evaluated in the present study, as our previous work had already confirmed excellent intra- and inter-observer reproducibility for this single-slice protocol [10]. MRI was chosen for its ability to assess skeletal muscle mass, intra-hepatic fat (PDFF), and liver stiffness (LS) in a single session, offering superior accuracy over CT or ultrasonography. Current AASLD guidance endorses imaging-based assessment of SP [11].

### 1.4 Outcome and Follow-up

Patients with advanced chronic liver disease (ACLD: LS $\geq$ 3 kPa) were followed from the date of baseline MRI to the date of liver-related death, liver transplantation, last clinic visit, or confirmation of survival. For patients who underwent liver transplantation, the date of transplant surgery was considered the censoring point. The primary outcome of the study was liver-related mortality, defined as death attributable to hepatic decompensation, variceal bleeding, spontaneous bacterial peritonitis, or hepatocellular carcinoma. Patient outcomes were initially reviewed through electronic medical records. For patients who were referred to other institutions, outcome data were obtained via interinstitutional communication. In cases where outcomes remained uncertain, direct contact with the patient or their family was attempted by phone. The median follow-up period was 34.0 months (interquartile range: 3–73 months). Patients with incomplete follow-up information were censored at the date of last confirmed contact.

### 1.5 Statistical Analysis

All statistical analyses were performed using JMP version 19.0.0 (SAS Institute Japan). Between-group differences were assessed using the chi-square test for categorical variables and the Mann–Whitney–Wilcoxon test for continuous variables. For comparisons among three or more groups, the Kruskal–Wallis test was used, followed by post hoc Steel–Dwass tests to identify significant pairwise differences.

Survival analysis in patients with ACLD was conducted using the Kaplan–Meier method, with differences between groups assessed by the log-rank test. A Cox proportional hazards model was employed to identify independent prognostic factors. For multivariate analysis, variables were initially selected using stepwise forward and backward methods, and a minimal set of clinically relevant factors was retained to construct the final model. Given the exploratory nature of univariate and subgroup comparisons, no correction for multiple testing was applied. In all analyses, a p-value of $< 0.05$ was considered statistically significant.

**Supplementary Table 1.** Univariable and multivariable predictors of Na <139 mEq/L in patients with chronic liver disease without hepatocellular carcinoma.

| Risk Factor | Univariable analysis | | Multivariable analysis | |
|---|---|---|---|---|
| | Odds ratio (95% CI) | P value | Odds ratio (95% CI) | P value |
| Sex (Male) | 1.57 (1.09-2.28) | 0.014 | **1.94 (1.12-3.36)** | **0.018** |
| Etiology (ArLD) | 3.27 (2.13-4.99) | <.001 | | |
| Total bilirubin (mg/dl) | 1.38 (1.18-1.65) | <.001 | | |
| γ-GTP (IU/L) | 1.01 (1.00-1.01) | 0.006 | | |
| Total Cholesterol (mg/dL) | 0.99 (0.98-0.99) | <.001 | | |
| Albumin (g/dl) | 0.29 (0.20-0.39) | <.001 | | |
| Leukocytes (/$mm^3$) | 1.01 (1.00-1.01) | <.001 | **1.01 (1.00-1.01)** | **0.007** |
| Hemoglobin (gldl) | 0.82 (0.75-0.89) | <.001 | | |
| Platelets (x$10^4$ /$mm^3$) | 0.96 (0.95-0.97) | <.001 | | |
| Prothrombin activity (%) | 0.96 (0.90-0.95) | <.001 | | |
| Ammonia (μg/dL) | 1.01 (1.01-1.02) | <.001 | 0.99 (0.98-1.00) | 0.271 |
| CRP (mg/dL) | 3.73 (2.37-6.19) | <.001 | | |
| LS (kPa) | 1.41 (1.30-1.54) | <.001 | **1.17 (1.02-1.34)** | **0.021** |
| Lymphocyte count (/$mm^3$) | 0.99 (0.98-0.99) | <.001 | 0.99 (0.99-1.00) | 0.271 |
| ALBI score | 3.90 (2.80-5.54) | <.001 | **2.55 (1.45-4.62)** | **0.001** |
| Presence of SP (n=210) | 2.08 (2.07-4.29) | <.001 | **2.09 (1.22-3.58)** | **0.008** |
| Presence of PHT (n=61) | 8.26 (4.78-14.57) | <.001 | | |

| | | |
|---|---|---|
| Use of diuretics (n=37) | 9.93 (4.96-2)0.92 | <.001 |

ALBI score, Albumin–Bilirubin score; ArLD, Alcohol-related Liver Disease; CI, Confidence Interval; CRP, C-reactive Protein; γ-GTP, γ-Glutamyl Transpeptidase; LS, Liver Stiffness; PHT, Portal Hypertension; SP, Sarcopenia.

**Supplementary Table 2.** Characteristics across four Liver–Muscle Phenotype (LMP) groups in non-HCC chronic liver disease.

| | **LMP 1 (n=591)** | **LMP 2 (n=158)** | **LMP 3 (n=24)** | **LMP 4 (n=65)** | P-value |
|---|---|---|---|---|---|
| Age (y) | 58±14 | 67±14* | 55±14† | 66±13*‡ | <.001 |
| Sex (Male%) | 57% | 65% | 76% | 71% | 0.035 |
| BMI (kg/m$^2$) | 25±4 | 22±3* | 27±5† | 23±6*‡ | <.001 |
| Obesity (BMI ≥25 kg/m$^2$) | 53% | 25% | 67% | 29% | <.001 |
| Sarcopenia (%) | 0 | 100% | 0 | 100% | - |
| Etiology: HBV | 20% | 25% | 14% | 2% | |
| HCV | 18% | 27% | 5% | 12% | |
| Alcohol | 8% | 18% | 43% | 58% | <.001 |
| MASLD | 41% | 12% | 33% | 12% | |
| Others | 13% | 18% | 5% | 17% | |
| Leucocytes (/mm$^3$) | 6056±1580 | 5511±1643* | 6507±3247 | 6842±5085 | <.001 |
| Hemoglobin (g/dl) | 14.2±1.7 | 13.6±1.8* | 12.6±3.0 | 10.8±1.8*‡ | <.001 |
| Platelets (x10$^4$/mm$^3$) | 22.1±6.0 | 19.5±6.5* | 14.2±7.3*† | 14.9±8.6*† | <.001 |
| Total bilirubin (mg/dL) | 0.9±0.4 | 0.9±0.6 | 3.1±2.6*† | 2.5±2.8*† | <.001 |

| | | | | | |
|---|---|---|---|---|---|
| AST (IU/L) | 37±31 | 37±31 | 227±500*† | 60±44*† | <.001 |
| ALT (IU/L) | 46±47 | 32±30* | 293±777† | 44±99 | <.001 |
| ALP (IU/L) | 251±118 | 277±159 | 480±246*† | 371±191*† | <.001 |
| γ-GTP (IU/L) | 82±117 | 108±220 | 354±457*† | 181±239*† | <.001 |
| Albumin (g/dl) | 4.3±0.3 | 4.2±0.3* | 3.1±0.5*† | 2.8±0.5*† | <.001 |
| Total Cholesterol (mg/dL) | 201±36 | 188±42* | 165±37* | 139±49*† | <.001 |
| Triglyceride (mg/dL) | 163±124 | 132±95* | 147±111* | 80±38*†‡ | <.001 |
| Prothrombin activity (%) | 100±14 | 98±14 | 65±18*† | 68±17*† | <.001 |
| LS (kPa) | 2.9±1.3 | 3.3±1.8* | 7.8±3.1*† | 7.1±3.2*† | <.001 |
| PDFF (%) | 9.5±7.9 | 5.6±5.3* | 5.6±6.1* | 4.6±6.2*† | <.001 |

Statistics are shown as the mean ± standard deviation, number or n (%). P-values represent comparisons among the four LMP groups by ANOVA or $\chi^2$ test, as appropriate. *: P<0.05 for LMP 1, †: P<0.05 for LMP 2, ‡: P<0.05 for LMP 3

Abbreviations: HR, hazard ratio; CI, confidence interval; ArLD, alcohol-related liver disease; ALT, alanine aminotransferase; ALP, alkaline phosphatase; CRP, C-reactive protein; LS, liver stiffness; PDFF, proton density fat fraction; ALBI, albumin–bilirubin score; MELD, model for end-stage liver disease score; ACLD, advanced chronic liver disease; HCC, hepatocellular carcinoma; LMP, liver–muscle phenotype.

**Supplementary Table 3.** Comparative Cox proportional hazards models (Model 1 and Model 2) for prediction of liver-related mortality in advanced chronic liver disease.

| | Model 1 | Model 2 |
|---|---|---|

| **Variables** | HR (95% CI) | P value | HR (95% CI) | P value |
|---|---|---|---|---|
| ALT (IU/L) | 0.99 (0.98–1.00) | 0.075 | 0.99 (0.97–1.00) | 0.059 |
| Trigliceride (mg/dL) | 0.99 (0.99–1.00) | 0.198 | 0.99 (0.99–1.00) | 0.182 |
| Sodium (mEq/L) | - | | - | |
| MELD Na | **1.08 (1.02–1.14)** | **0.010** | - | |
| MELD 3.0 | - | | **1.11 (1.05–1.18)** | **0.001** |
| Presence of HCC | **5.26 (2.84–9.72)** | **<0.001** | **5.40 (2.92–9.99)** | **<.001** |
| LMP 4 (vs. LMP 1-3) | **5.29 (2.53–9.78)** | **<0.001** | **4.16 (2.05–8.45)** | **<.001** |

**Statistics are expressed as hazard ratios (HRs) with 95% confidence intervals (CIs).**

**Model 1** included ALT, triglyceride, serum sodium, MELD-Na, presence of HCC, and LMP classification.

**Model 2** included ALT, triglyceride, serum sodium, MELD 3.0, presence of HCC, and LMP classification.

**Abbreviations:** HR, hazard ratio; CI, confidence interval; ALT, alanine aminotransferase; MELD, model for end-stage liver disease; HCC, hepatocellular carcinoma; LMP, liver–muscle phenotype.

**Supplementary Table 4.** Comparative clinical and laboratory characteristics between Liver–Muscle Phenotype (LMP) 3 and 4 in advanced chronic liver disease.

| | **LMP 3 (n=24)** | **LMP 4 (n=65)** | P value |
|---|---|---|---|
| Age (y) | 58 (48, 69) | 67 (57, 79) | **0.012** |
| Sex (Male/Female) | 19/5 | 46/19 | 0.420 |
| BMI (kg/m$^2$) | 26 (24, 30) | 23 (21, 26) | **<.001** |
| Subcutaneous fat area (cm$^2$) | 99 (68, 169) | 58 (42, 82) | **<.001** |
| Obesity (BMI ≥25 kg/m$^2$) | 15 (63%) | 20 (31%) | **<.001** |
| Etiology (ALD) | 7 (29%) | 34 (52%) | **0.049** |
| Presence of HCC | 7 (29%) | 16 (25%) | 0.666 |
| Decompensated ACLD | 12 (58%) | 53 (82%) | **0.029** |
| Total bilirubin (mg/dL) | 1.8 (1.2, 4.1) | 1.5 (1.0, 2.5) | 0.244 |
| AST (IU/L) | 49 (39, 77) | 54 (34 82) | 0.886 |
| ALT (IU/L) | 28 (24, 60) | 28 (16, 42) | 0.106 |
| ALP (IU/L) | 439 (302, 600) | 356 (269, 512) | 0.256 |
| γ-GTP (IU/L) | 150 (49, 288) | 95 (36, 238) | 0.266 |
| PT (INR) | 1.3 (1.2, 1.5) | 1.2 (1.1, 1.4) | 0.360 |
| BUN (mg/dL) | 11 (7, 16) | 16 (11, 23) | **0.009** |
| Creatinine (mg/dL) | 0.7 (0.6, 0.9) | 0.9 (0.6, 1.0) | 0.135 |
| Sodium (mEq/L) | 141 (137, 143) | 137 (136, 140) | **0.004** |
| Albumin (g/dl) | 3.2 (2.9, 3.5) | 2.9 (2.6, 3.2) | **0.003** |
| Total Cholesterol (mg/dL) | 156 (133, 178) | 134 (110, 157) | **0.030** |
| Triglyceride (mg/dL) | 104 (67, 188) | 71 (58, 104) | **0.026** |

| | | | |
|---|---|---|---|
| Hemoglobin (g/dl) | 14 (10, 15) | 11 (10, 12) | **0.003** |
| Lymphopenia (<1500/mm$^3$) | 10 (42%) | 51 (79%) | **0.001** |
| Ammonia (μg/dL) | 80 (50, 117) | 51 (35, 96) | 0.131 |
| CRP (mg/dL) | 0.5 (0.1, 1.0) | 0.9 (0.3, 2.3) | **0.009** |
| PDFF (%) | 4.4 (2.7, 6.7) | 2.2 (1.6, 3.7) | **0.003** |
| MRE-LS (kPa) | 7.7 (5.6, 9.6) | 7.0 (5.5, 8.9) | 0.482 |
| ALBI score | -1.6 (-1.9, -1.4) | -1.5 (-1.8, -1.1) | 0.132 |
| MELD score | 13 (9, 16) | 12 (9, 18) | 0.580 |
| MELD Na | 13 (9, 18) | 13 (9, 18) | 0.788 |
| MELD 3.0 | 13 (10, 19) | 14 (10, 18) | 0.656 |

**Statistics are shown as the median (interquartile range) or number (%).**

P-values represent comparisons between LMP 3 and LMP 4 groups by Mann–Whitney U test or $\chi^2$ test, as appropriate.

**Abbreviations:**

ALD, alcohol-related liver disease; ACLD, advanced chronic liver disease; ALBI, albumin–bilirubin score; ALT, alanine aminotransferase; AST, aspartate aminotransferase; ALP, alkaline phosphatase;

BUN, blood urea nitrogen; CRP, C-reactive protein; γ-GTP, γ-glutamyl transpeptidase; PT, prothrombin time; INR, international normalized ratio; PDFF, proton density fat fraction; MRE-LS, magnetic resonance elastography–liver stiffness; MELD, model for end-stage liver disease score; MELD-Na, MELD–sodium score; HCC, hepatocellular carcinoma; LMP, liver–muscle phenotype.

**Supplementary Figures.**

**Supplementary Fig 1. Flowchart showing inclusion and exclusion of patients in this study.** ACLD, advanced chronic liver disease; HCC, Hepatocellular carcinoma; MRI, magnetic resonance imaging; MRE, magnetic resonance elastography.

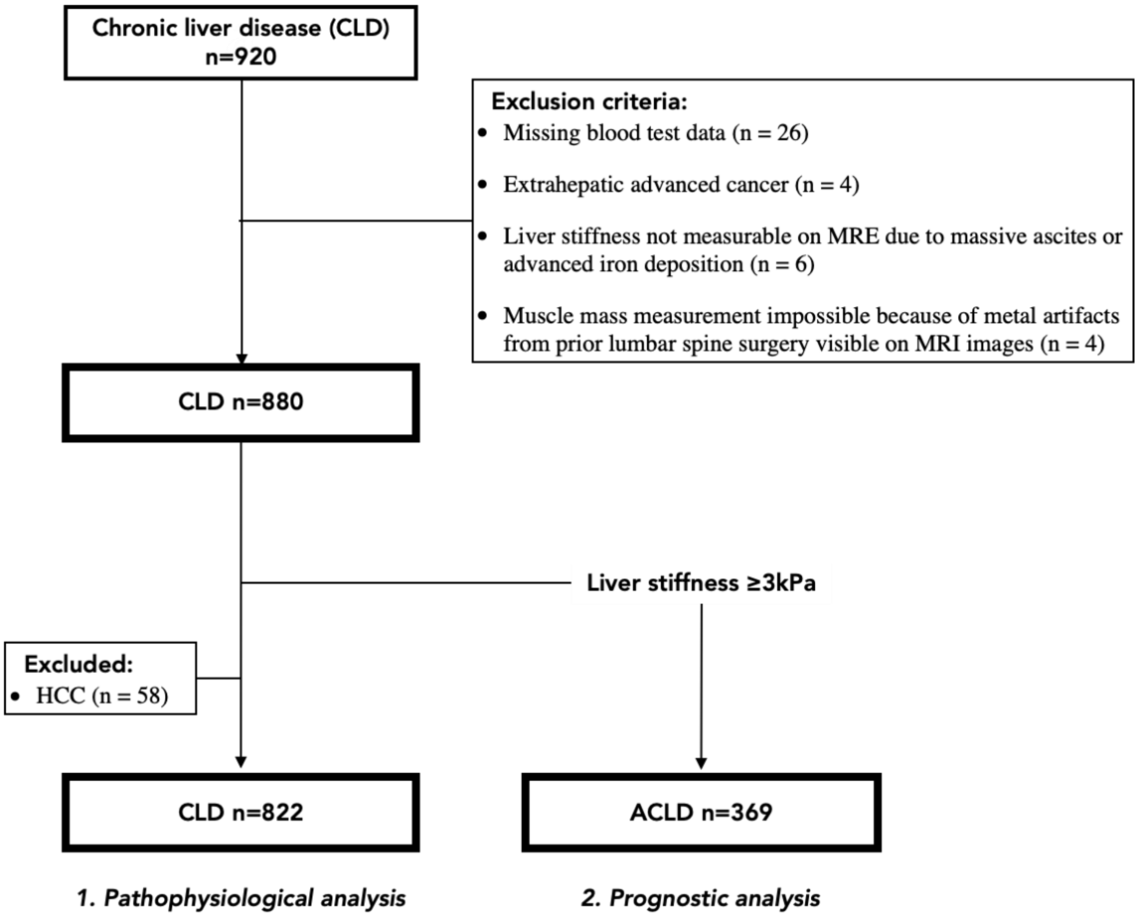

**Supplementary Figure 2.** Sex-specific analyses of serum sodium–age relationships and the predictive cutoff of Na for sarcopenia.

(A) No sex-related difference in serum sodium–age relationships

(B) Serum Na cutoff predicting sarcopenia was 138 mEq/L in both sexes

**(A)**

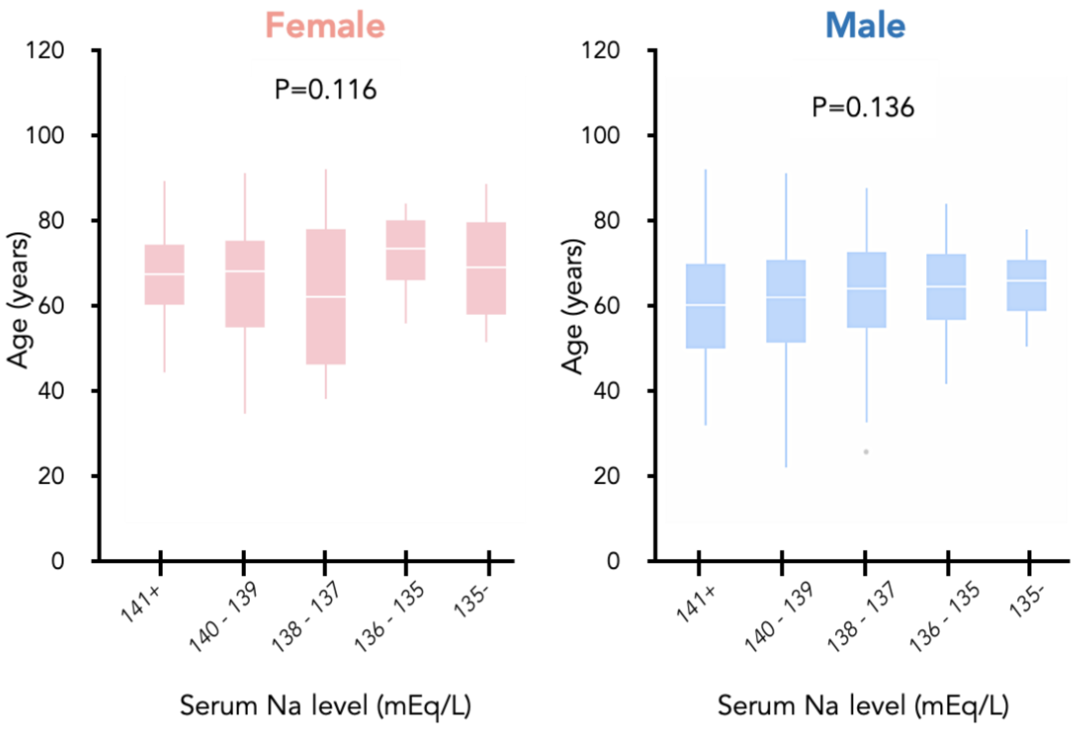

**(B)**

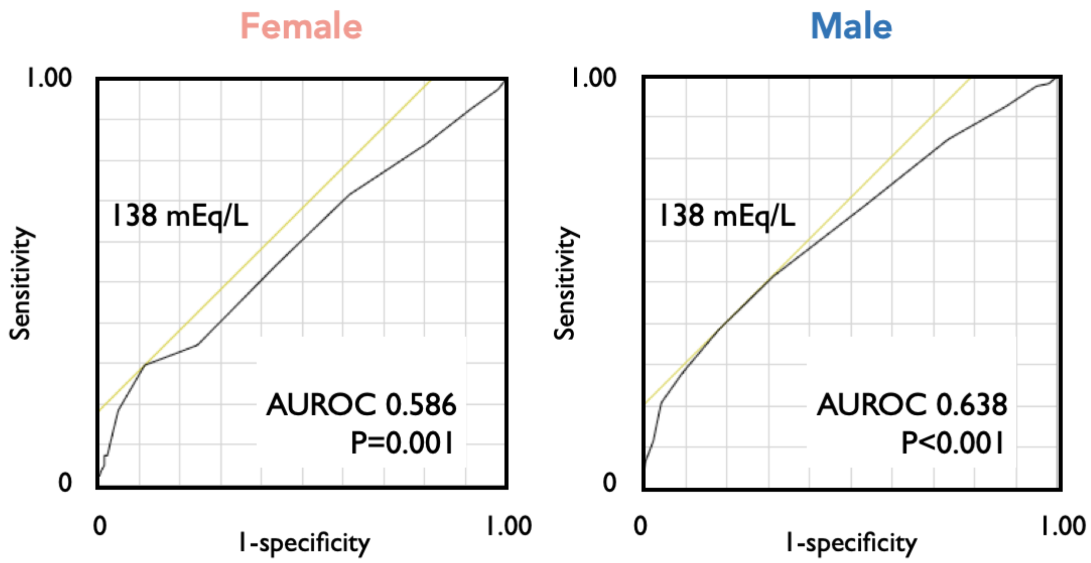


**Supplementary Figure 3.**

**(A)** ROC analysis identifying the MRE-based liver stiffness cutoff corresponding to the ALBI −2.16 threshold used in the Figure 3 decision tree.

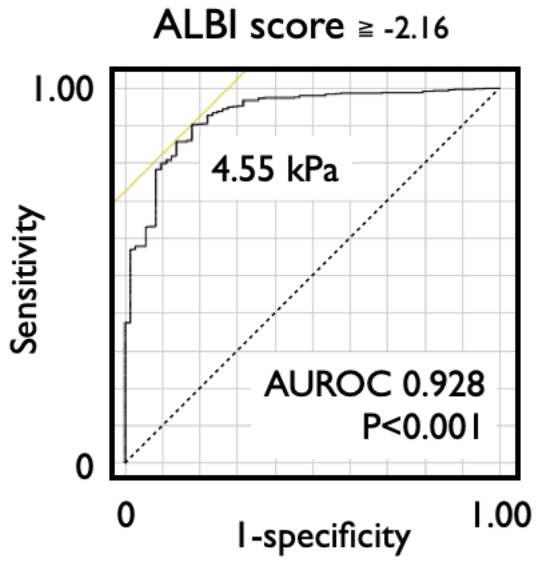

**(B)** Extended decision tree analysis incorporating diuretic use into the Figure 3 model.

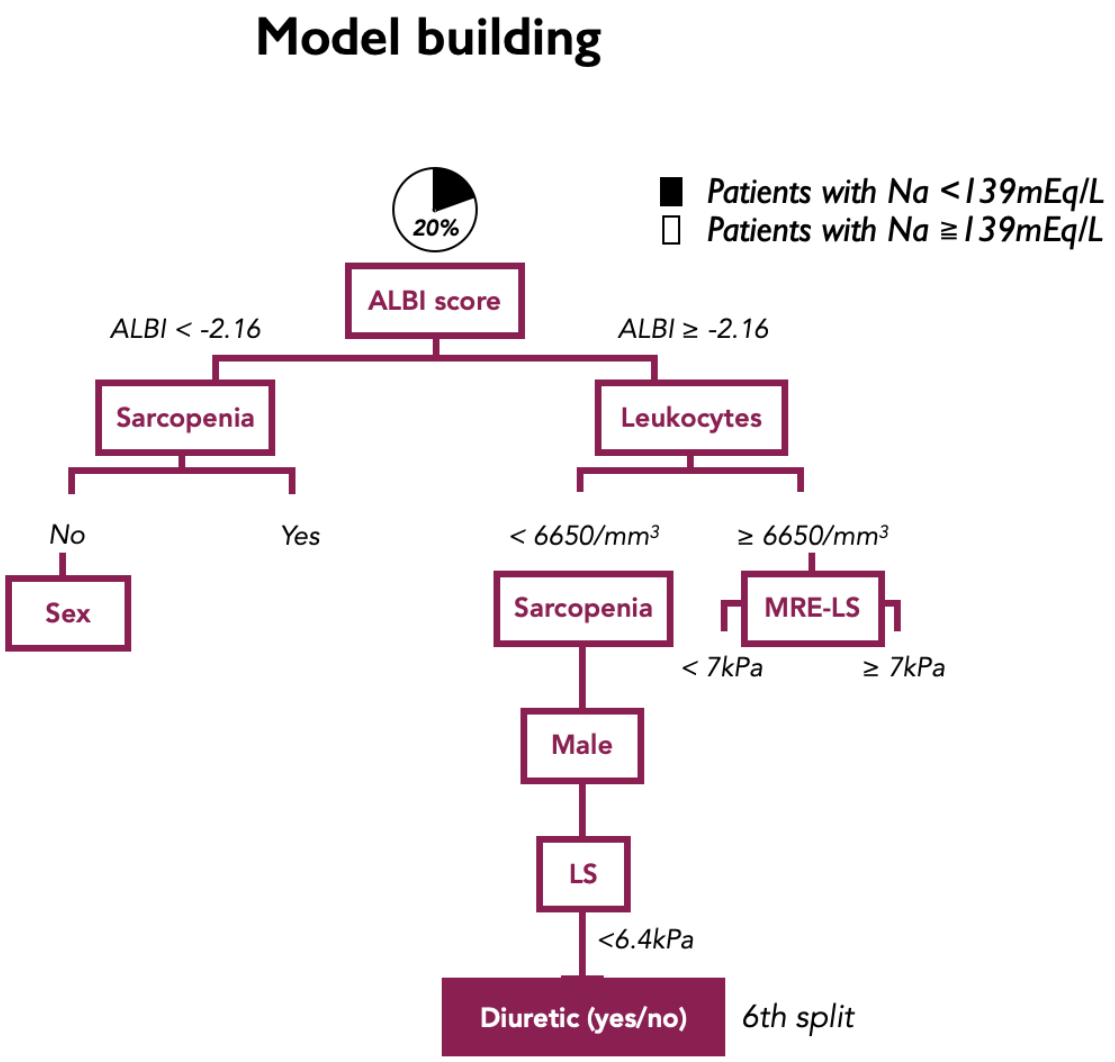


- Cut-offs auto-selected by CART algorithm
- %: prevalence of patients with sodium <139mEq/L in that node

**Supplementary Figure 4.** New-onset ascites in compensated ACLD according to serum sodium levels (<139 vs ≥139 mEq/L); adjusted HR 3.07 (P=0.019).

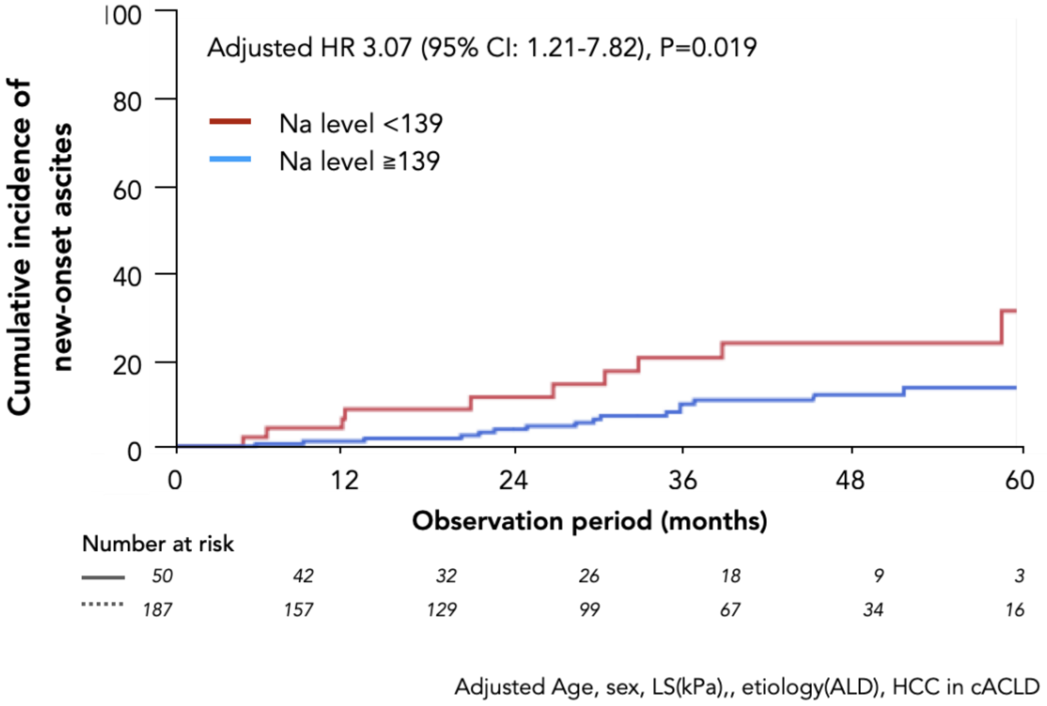

**Supplementary Figure 5. Adjusted risk of clinical deterioration according to Liver–Muscle Phenotype**

Adjusted hazard ratios for clinical deterioration using LMP1 as the reference group were 3.78 (95% CI 1.31–10.88, P=0.014) for LMP2, 5.67 (95% CI 1.68–19.07, P=0.005) for LMP3, and 15.98 (95% CI 5.37–47.53, P<0.001) for LMP4. The model was adjusted for age, sex, liver stiffness (kPa), HCC, etiology (alcohol-related liver disease), and diuretic use.

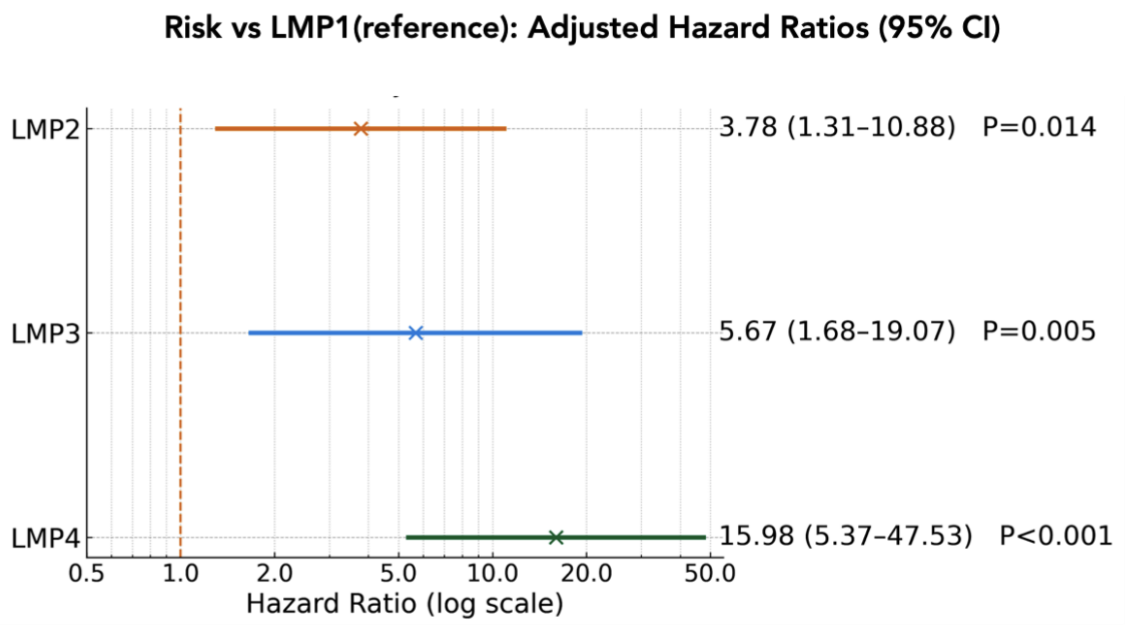


Adjusted for age, sex, liver stiffness (kPa), HCC, etiology(ArLD), and diuretic use.
Log scale; dashed line indicates HR=1.

**Supplementary Figure 6.** Overall survival according to serum Na level categories in chronic liver disease. Survival differed significantly across serum sodium strata (≥141, 140–139, 138–137, 136–135, and <135 mEq/L), with patients below 139 mEq/L demonstrating worse outcomes (P<0.001). Numbers at risk are shown below the Kaplan–Meier curves.

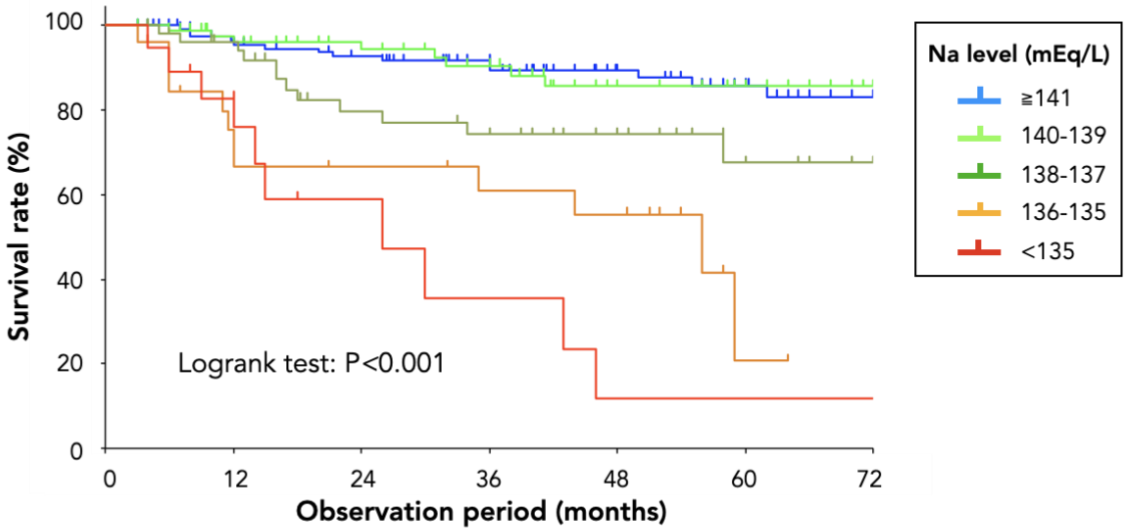


Number at risk

| | | | | | | | |
|---|---|---|---|---|---|---|---|
| ≧141 | 150 | 112 | 99 | 86 | 59 | 41 | 14 |
| 140-139 | 106 | 83 | 67 | 55 | 34 | 27 | 15 |
| 138-137 | 61 | 48 | 33 | 29 | 20 | 12 | 5 |
| 136-135 | 30 | 20 | 16 | 13 | 12 | 3 | 0 |
| <135 | 22 | 14 | 7 | 5 | 3 | 3 | 3 |

**Supplementary Figure 7.** Survival analyses showing poorer prognosis in patients with serum Na <139 mEq/L across ACLD, non-HCC, and HCC groups (Log-rank P<0.001).

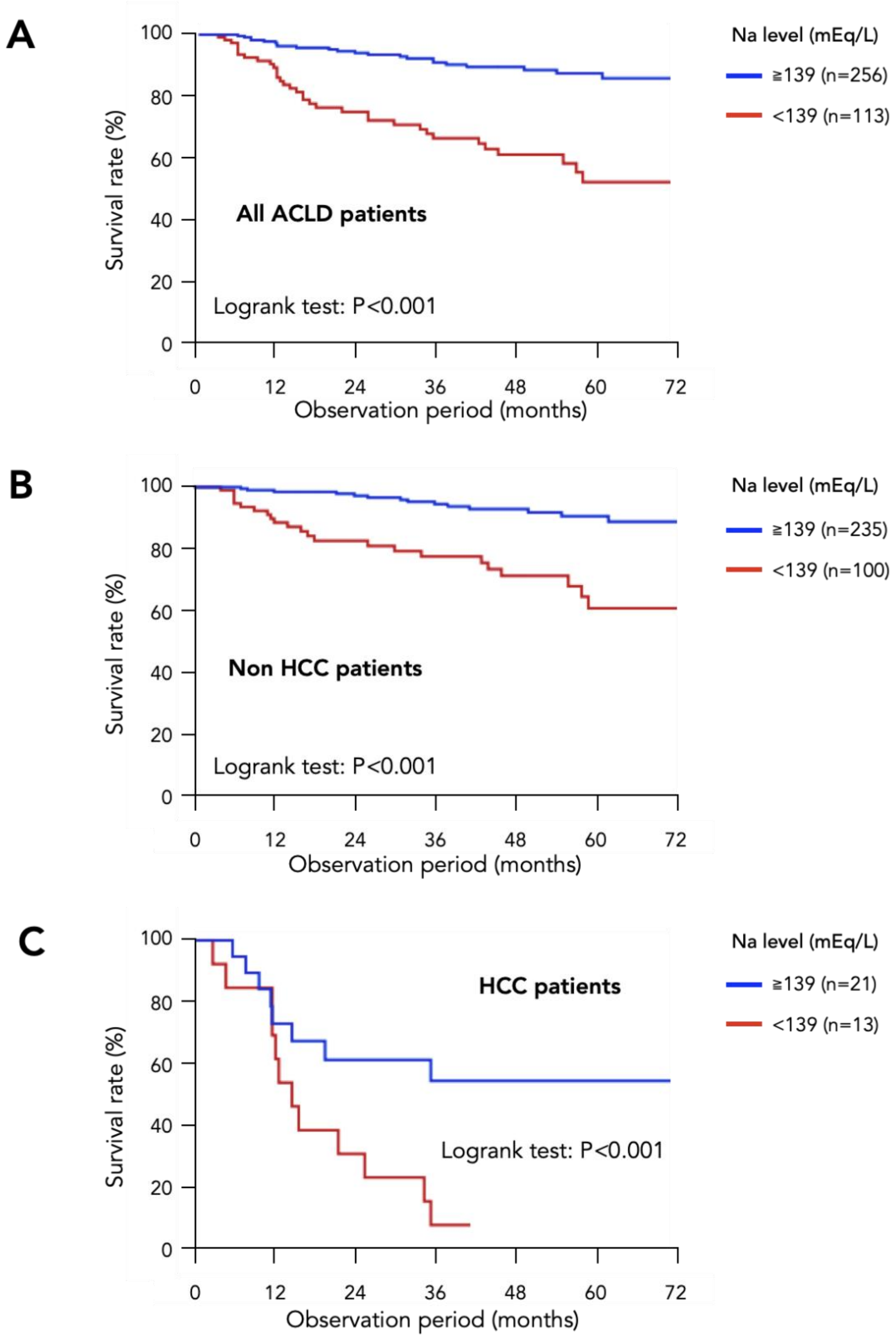

**Supplementary Figure 8.** Correlation between hemoglobin and total bilirubin in non-ACLD patients (LS <3 kPa) without alcohol-related liver disease (ρ=0.287, P<0.001; n=456).

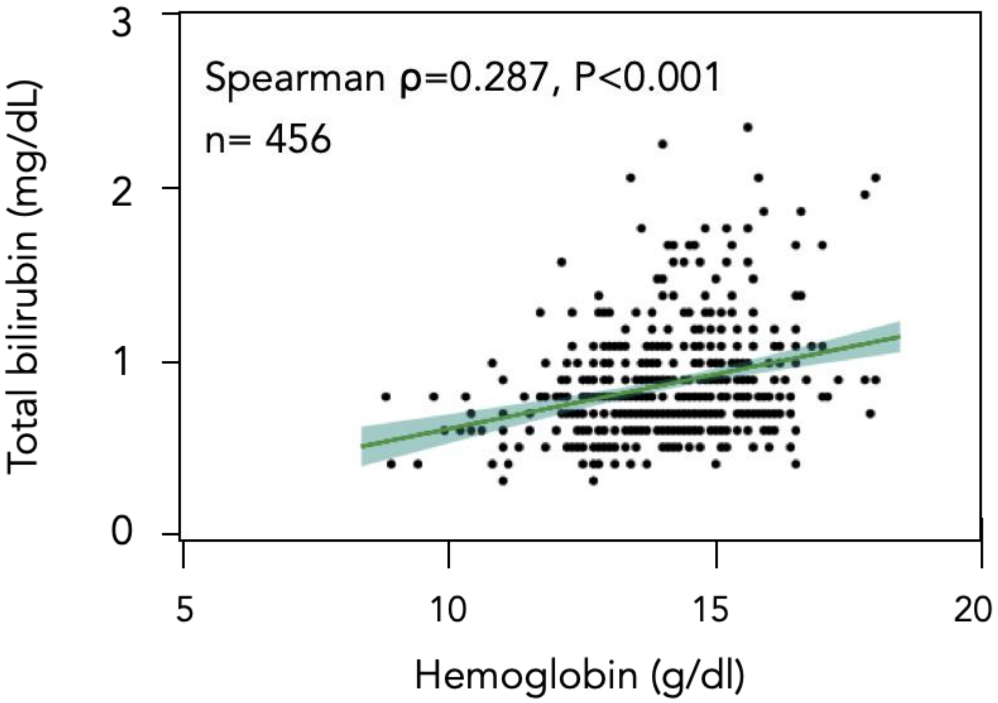
